# Multivariate age-related variations in quantitative MRI maps: Widespread age-related differences revisited

**DOI:** 10.1101/2023.10.19.23297253

**Authors:** Soodeh Moallemian, Christine Bastin, Martina F. Callaghan, Christophe Phillips

## Abstract

This study applied multivariate ANOVA to investigate age-related microstructural changes in the brain tissues driven primarily by myelin, iron, and water content, as observed in MRI (semi-)quantitative R1, R2*, MTsat and PD maps. This is effectively a re-analysis of the data analyzed in a univariate way by Callaghan et al., 2014. Voxel-wise analyses were performed on gray matter (GM) and white matter (WM), in addition to region of interest (ROI) analyses. The multivariate approach identified brain regions showing coordinated alterations in multiple tissue properties and demonstrated bidirectional correlations between age and all examined modalities in various brain regions, including the caudate nucleus, putamen, insula, cerebellum, lingual gyri, hippocampus, and olfactory bulb. The multivariate model was more sensitive than univariate analyses, as evidenced by detecting a larger number of significant voxels within clusters in the supplementary motor area, frontal cortex, hippocampus, amygdala, occipital cortex, and cerebellum bilaterally. Though when cross validating the results by splitting the data into 2 subsets, sensitivity is strongly reduced, even more so for the multivariate approach. The examination of normalized, smoothed, and z-transformed maps within the ROIs revealed concurrent age-dependent alterations in myelin, iron, and water content. These findings contribute to our understanding of age-related brain differences and provide insights into the underlying mechanisms of aging. The study emphasizes the importance of multivariate analysis for detecting subtle microstructural changes associated with aging when dealing with multiple quantitative MRI parameter maps.

## 1. Introduction

Aging is an inevitable part of our lifecycle associated with the physical deterioration of different organs. Various hallmarks of aging have been identified over the past years that associate with neurodegenerative pathological changes in the brain, making age a primary risk factor for most neurodegenerative diseases, including Alzheimer’s disease (AD), Parkinson’s disease (PD), and frontotemporal lobar dementia (FTD) (Azam et al., 2021; Jeremic et al., 2021).

Quantitative MR imaging (qMRI) enables us to extract sensitive and specific information about the microstructural properties of the brain tissue *in-vivo*, such as axon, myelin, iron, and water concentration (Weiskopf et al., 2021). The estimation of (semi-)quantitative metrics normally includes effective longitudinal relaxation rate (R1), which is sensitive to iron, myelin and water content, transverse relaxation rates (R2*), which is primarily sensitive to iron, proton density (PD) indicative of free water content, and magnetization transfer saturation (MTsat) associated with macromolecular content, predominantly myelin (Tabelow et al., 2019a; Taubert et al., 2020a).

Many studies of aging and neurodegenerative diseases focus on alterations in the nervous system, such as (de-)myelination or iron accumulation (Moallemian et al., 2023; Peters, 2002; Tian et al., 2022). Callaghan et al. (2014) investigated age-related differences of biologically relevant *in-vivo* measures over the course of normal aging using quantitative multiparameter mapping (MPM) and showed significant demyelination in white matter (WM), concurrent with an increase in iron levels in the basal ganglia, red nucleus, and extensive cortical regions, but decreases along the superior occipitofrontal fascicle and optic radiation (Callaghan et al., 2014). Steiger et al. (2016) investigated the difference in iron and myelin levels between two groups of young and older participants using MPM qMRI, and showed that age-related higher levels of iron are accompanied by a negative correlation of iron and myelin in the ventral striatum (Steiger et al., 2016). Although demyelination primarily affects the WM of the brain, recent research shows that it can also occur in gray matter (GM), which is made up of the cell bodies of neurons. Studies have shown that gray matter myelination can occur during development, particularly in the prefrontal cortex, and may continue throughout late adulthood in response to learning and experience (Fields, 2008; Timmler and Simons, 2019). Khattar and colleagues assessed the association of myelination and iron accumulation in the brain of a cohort of 21-94 year-old healthy controls and found a negative correlation between whole brain myelin water fraction and iron content in most brain regions; they also highlighted that the myelination continues until middle age in the brain (Khattar et al., 2021). Another study reported robust evidence for spatial overlap between volume, myelination, and iron decomposition changes in aging that affect predominantly motor and executive networks under a modified normal probability curve approach from the Permutation Analysis of Linear Models (PALM) toolbox (Taubert et al., 2020b; Winkler et al., 2016, 2014).

In this technical note, we re-analyze the data from (Callaghan et al., 2014), now available as an open dataset on OpenNeuro (doi:10.18112/openneuro.ds005851.v1.0.0), with a multivariate statistical modeling approach as implemented in the MSPM toolbox (Gyger et al., 2021), available on GitHub (https://github.com/LREN-CHUV/MSPM), with the code to reproduce the results also available on GitHub (https://github.com/CyclotronResearchCentre/qMRI_MSPM_AgeFx). While the dataset examined here has been previously characterized using univariate and region-specific approaches, the present study adopts a multivariate framework to assess how multiple imaging modalities jointly relate to aging. This integrative perspective enables the examination of across-modality effects that may not be captured by traditional univariate analyses. Moreover, we performed a simple cross-validation, by splitting the original data into 2 subsets and repeating the analysis on these.

## 2. Method

### 2.1. Participants and spatial preprocessing

We took advantage of the processed data from (Callaghan et al., 2014; Callaghan and Phillips, 2025; Phillips et al., 2025) which include 138 healthy participants aged 19-75 years (35.5% male, mean = 46.64, s.d = 21). See in the “Supplementary material” for a full description of the acquisition, as provided in (Callaghan et al., 2014).

(Semi-)quantitative multiparameter maps (R1, R2*, PD, and MTsat) were reconstructed with the VBQ toolbox, a preliminary version of the hMRI toolbox (Draganski et al., 2011; Tabelow et al., 2019b). Processing steps included segmentation using the “unified segmentation” approach (Ashburner and Friston, 2005), and diffeomorphic morphing to MNI space using DARTEL (Ashburner, 2007). Tissue-weighted smoothing (for GM and WM separately) with a 3mm FHWM isotropic kernel was applied to account for residual misalignment while preserving the quantitative nature of the data. These steps produced 8 smoothed normalized qMRI maps, one for each parameter (R1, R2*, PD, and MTsat) and tissue class (GM and WM). Finally, group-level GM and WM masks were created to be further used as exclusive masks in the statistical analysis. For full details of the processing steps, see (Callaghan et al., 2014; Callaghan and Phillips, 2025).

### 2.2. Maps normalization and univariate GLM (uGLM) analyses

In this re-analysis, as recommended for MSPM (Gyger et al., 2021), the 8 resulting sets of qMRI maps were z-transformed per modality and across subjects to ensure comparability between maps, utilizing the mean and variance over each voxel. This procedure ensured comparability of different modalities for our multivariate analysis.

The univariate general linear models (uGLMs) were performed on each set of z-transformed maps using age, gender, total intracranial volume (TIV), and scanner as regressors. After statistical inference, on the effect of age, the 4 thresholded statistical maps are aggregated by taking their union, which we will refer to as the “union of uGLMs” (UuGLMs).

### 2.3. Multivariate GLM (mGLM)

While a standard multivariate regression model is useful for predicting the values of the dependent variables, it doesn’t directly address the question of how the entire set of dependent variables relates to the entire set of independent variables. In the context of this study, it is well-established that the brain undergoes simultaneous changes as we age. Therefore, a single independent variable may not adequately predict a single dependent variable; instead, the relationship might be more complex, where a combination of predictors may explain variance across multiple outcomes. Hence, a multivariate approach appears as a method of choice.

Here, we describe the multivariate GLM (mGLM) and its principles, including model and inference, as implemented in the MSPM toolbox are described here after (Gyger et al., 2021). For a more in depth description, refer to the original publication (Gyger et al., 2021) and the Supplementary material.

The multivariate GLM (mGLM) is specified using the design matrices of the 4 univariate models and the multivariate observations at each voxel as *Y* = *XB* + *E*, where *Y*_138×4_ = [*Y*_1_, *Y*_2_, *Y*_3_, *Y*_4_] is the multi-modal data matrix, each row of *Y* represents one participant, and each column represents one map; and *X*_138×5_ = [*X*_1_, *X*_2_, *X*_3_, *X*_4_, *X*_5_] is the design matrix, the first column represents the mean over subjects, the rest of the columns represent mean-centered regressors (age, TIV, gender, scanner) respectively. Thus, *B* is a 5 × 4 matrix of regression coefficients estimated using an ordinary least-square method, as 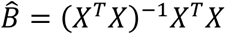 and 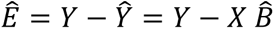 is the residual matrix of size 138 × 4. Importantly, the residuals are estimated on a per-voxel basis that allows for a straightforward determination of a unique covariance structure for each voxel. This feature is a significant advantage of mass multivariate approaches when dealing with dependent neuroimaging data. However, it is important to note that in this framework, the assumption of normality for the residuals implies that the covariance structure is assumed to be the same across different groups or conditions. There is an assumed degree of correlation between the columns of *Y*, this correlation is expressed by estimation of variance-covariance matrix 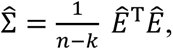 where n is the number of subjects and k is number of covariates.

#### Hypothesis Testing

Hypothesis testing in mGLM, relies on testing the linear contrast *CBL* = 0. This extension of the univariate scheme combines standard hypotheses on the rows of matrix *B*, i.e. on the regressor, represented by matrix *C*, with hypotheses on the columns of *B*, i.e. on the different modalities, represented by matrix *L*. In the context of multivariate ANOVA (MANOVA) models, contrasts of main effects and interactions are formulated by setting *L* = *I*_*t*_, the *t* × *t* identity matrix, as the dependent variables in multimodal neuroimaging applications are not assumed to be directly proportional. This method is the most suitable for conducting hypothesis testing on multimodal neuroimaging applications.

#### Test Statistics in mGLM

The hypothesis *H*_0_: *CBL* = 0 can be tested to assess the different potential co-occurrence of change between the modalities through Wilks’ lambda (Wilks, 1932) Λ test statistic, which has an approximate F distribution. To test the joint effect on the 4 (semi-)quantitative maps in a specific tissue type, *L*_4×4_is defined as an identity matrix corresponding to the number of dependent variables ((semi-)quantitative maps). As explained before, each column of *L* will perform a univariate analysis on each column of *B*. Here, *C* was defined as [0 1 0 0 0] to only see the correlation between age and maps. In this case, Λ has an exact F distribution with *a* = 4 and *b* = 130 degrees of freedom.

#### Canonical Correlation Analysis

Canonical vectors are calculated under the assumption that matrix *L* involves multiple dependent variables (*l* > 1), to extract the contribution of each dependent variable to the test statistics Λ. The number of possible canonical variates is equal to the number of variables in the smaller of the two sets of dependent and independent variables (in our case, 4). The canonical variates express the relative contribution of each variable to the tested effect.

### 2.4. Statistical inference and multiple comparison

All statistical analyses were performed on standardized z-transformed data. To assess the correlation between the individual tissue property maps and age, an F-test was performed on each map at voxel-level over GM and WM separately. All univariate statistical analyses were performed under the general linear model framework in SPM12, considering two p-values, .05 and .0125, family-wise error rate corrected (FWER) thresholds. The latter threshold accounts for the fact that for both tissue classes, 4 similar inferences are performed (one per parametric map); thus, applying a Bonferroni correction, the applied threshold is divided by 4, i.e., .0125 = .05/4. The former threshold, on the other hand, aims at replicating the previously published results (Callaghan et al., 2014).

With the multivariate GLM, one could simply rely on a .05 FWER threshold, as only one inference is performed for all 4 maps together.

### 2.5. Univariate and multivariate model evaluation

The results from the univariate and multivariate models are first compared in terms of number of significant voxels and clusters, as obtained from the thresholded statistical maps. Furthermore, we calculated Cohen’s Kappa Κ to assess the match between thresholded statistical maps because it provides a fair evaluation of “inter-rater reliability” (here the mGLM vs UuGLM), while accounting for the overall sparsity of significant voxels (Cohen, 1960). Moreover, to assess the stability and robustness of the statistical analyses, with both univariate and multivariate, we further conducted a split-half validation analysis. The dataset was randomly divided into two independent subsets, and the full analytical pipeline was repeated separately within each subset. Consistency of the results across splits was evaluated to determine the reliability of the observed multivariate patterns. Detailed results from this validation are provided in the Supplementary Materials (Tables S1–S4 and Figures S3–S5).

### 2.6. ROI analyses

The age-related parameter differences in selected regions, including caudate, cerebellum, hippocampus, middle frontal gyrus, pallidum, putamen, superior motor cortex, heschl and precentral gyri, and thalamus, will be further described. ROIs were selected based on identified significant areas in our mGLM results in (Callaghan et al., 2014). The ROIs were defined according to the Neuro-morphometrics AAL3 atlas (Rolls et al., 2020). As this atlas contains multiple subregions of thalamus and cerebellum, to create their ROI masks, we took the union of all thalamus and cerebellum subregions. For each participant and (semi-)quantitative map, the measure from each voxel in an ROI was extracted. The regressors of no interest (gender, TIV and scanner) were regressed out, and for each selected ROI, the relation between age and median value across subjects will be examined post hoc.

## 3. Results

The analyses were conducted on both gray matter (GM) and white matter (WM). Consequently, explicit non-overlapping masks for GM and WM were applied to each analysis. Since each parametric map has its specific unit, such as Hertz for R1 and R2* maps and p.u. for MTsat maps, their values cannot be directly compared, and the data were z-normalized (as indicated in Section 2.2).

### 3.1. uGLM vs mGLM: Voxel level analyses

The individual GM and WM analyses on R1, R2*, PD, and MTsat maps at a corrected threshold of p<.05 FWER, concur with those in (Callaghan et al., 2014). The statistical parametric maps for age-related changes in the microstructure of the brain in GM and WM thresholded at a standard p<.05 FWER are depicted in Figure S1 and Figure S2 of the supplementary data. The statistical parametric maps for the same analyses thresholded at a corrected p<.0125 FWER are presented in Figures ***Figure 1****-4* and ***Figure 5***-*8* for GM and WM, respectively. Voxel-wise mGLM results presented in ***Figure 9*** indicate bidirectional correlation between all modalities and age at p<.05 FWER corrected. The correlation is observed bilaterally in caudate nucleus, putamen, insula, cerebellum, lingual gyri, hippocampus, and olfactory bulb which matches the regions observed with the uGLMs.

**Figure 1.**
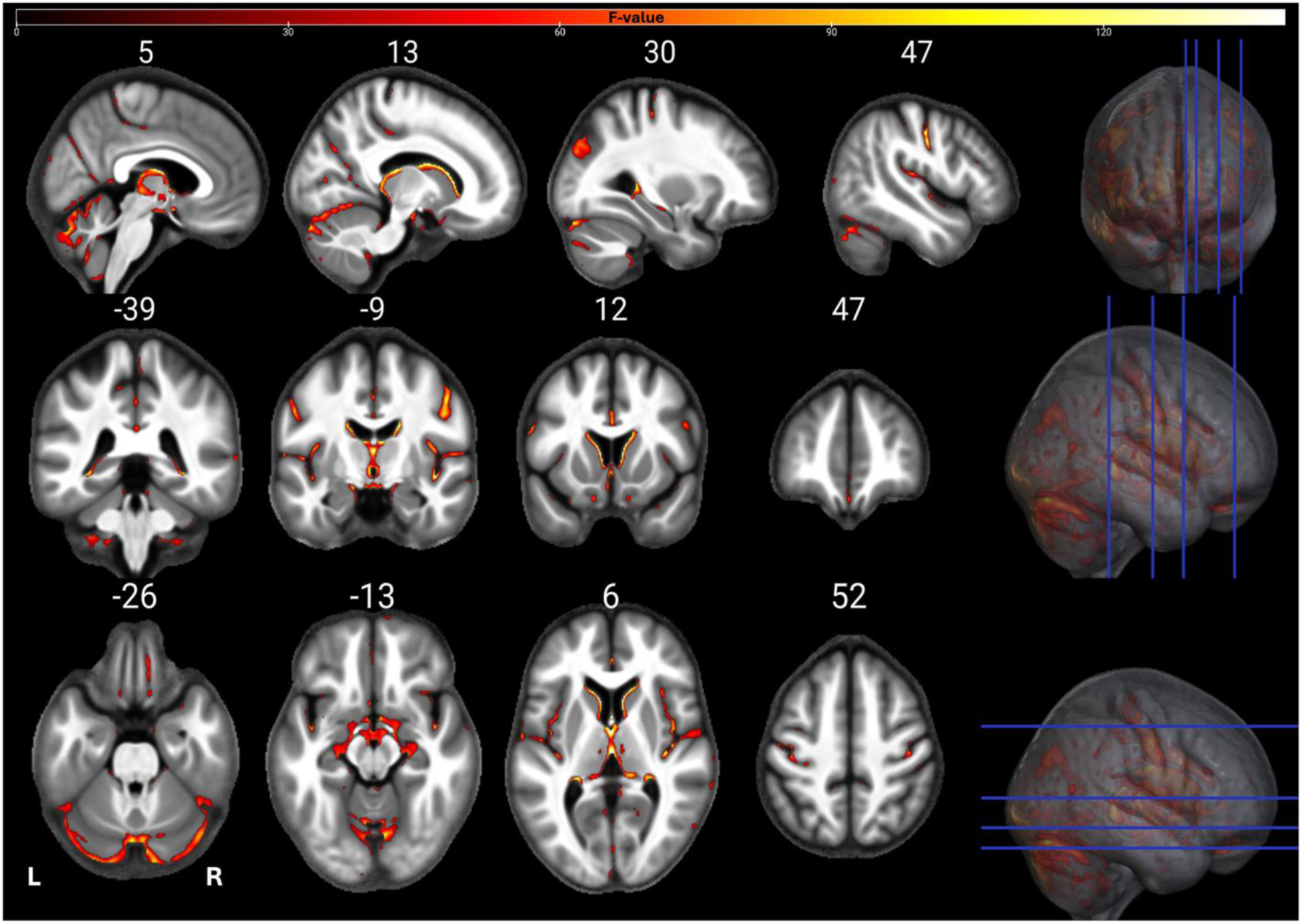
Statistical parametric maps of uGLM *MTsat* in GM; showing all the voxels with significant correlation with age, as detected by uGLMs for MTsat maps. The F-tests were thresholded at p<.0125 FWER corrected at voxel-level. The SPMs were overlayed on the mean MTsat map for the cohort in the MNI space. Abbreviation: GLM, general linear model; uGLM, univariate GLM; GM, gray matter; FWER, family-wise error rate; SPM, statistical parametric map.

**Figure 2.**
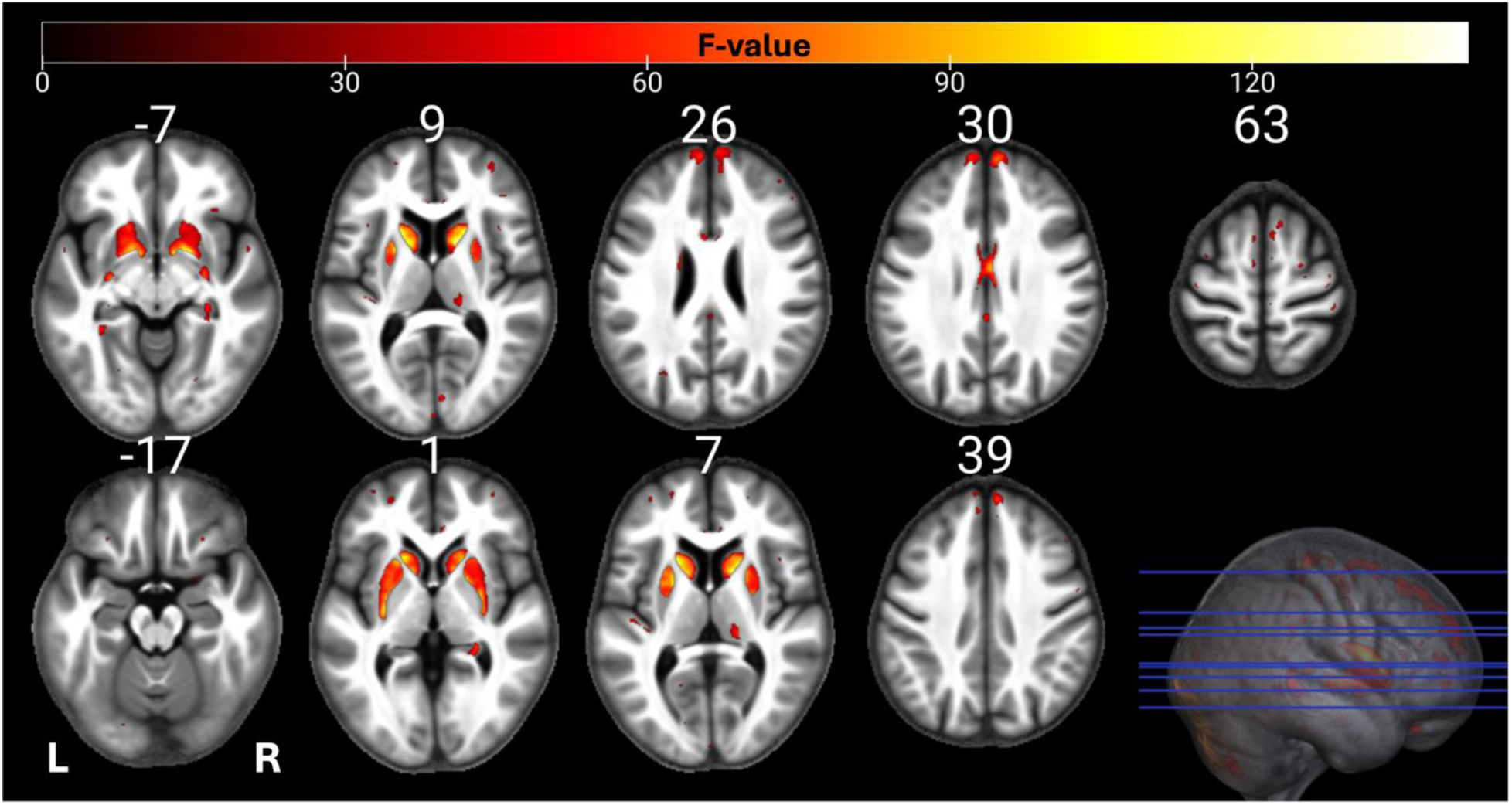
Statistical parametric maps of uGLM PD in GM; showing all the voxels with significant correlation with age, as detected by uGLMs for PD maps. The F-tests were thresholded at p<.0125 FWER corrected at voxel-level. The SPMs were overlayed on the mean MTsat map for the cohort in the MNI space. Abbreviation: GLM, general linear model; uGLM, univariate GLM; GM, gray matter; FWER, family-wise error rate; SPM, statistical parametric map.

**Figure 3.**
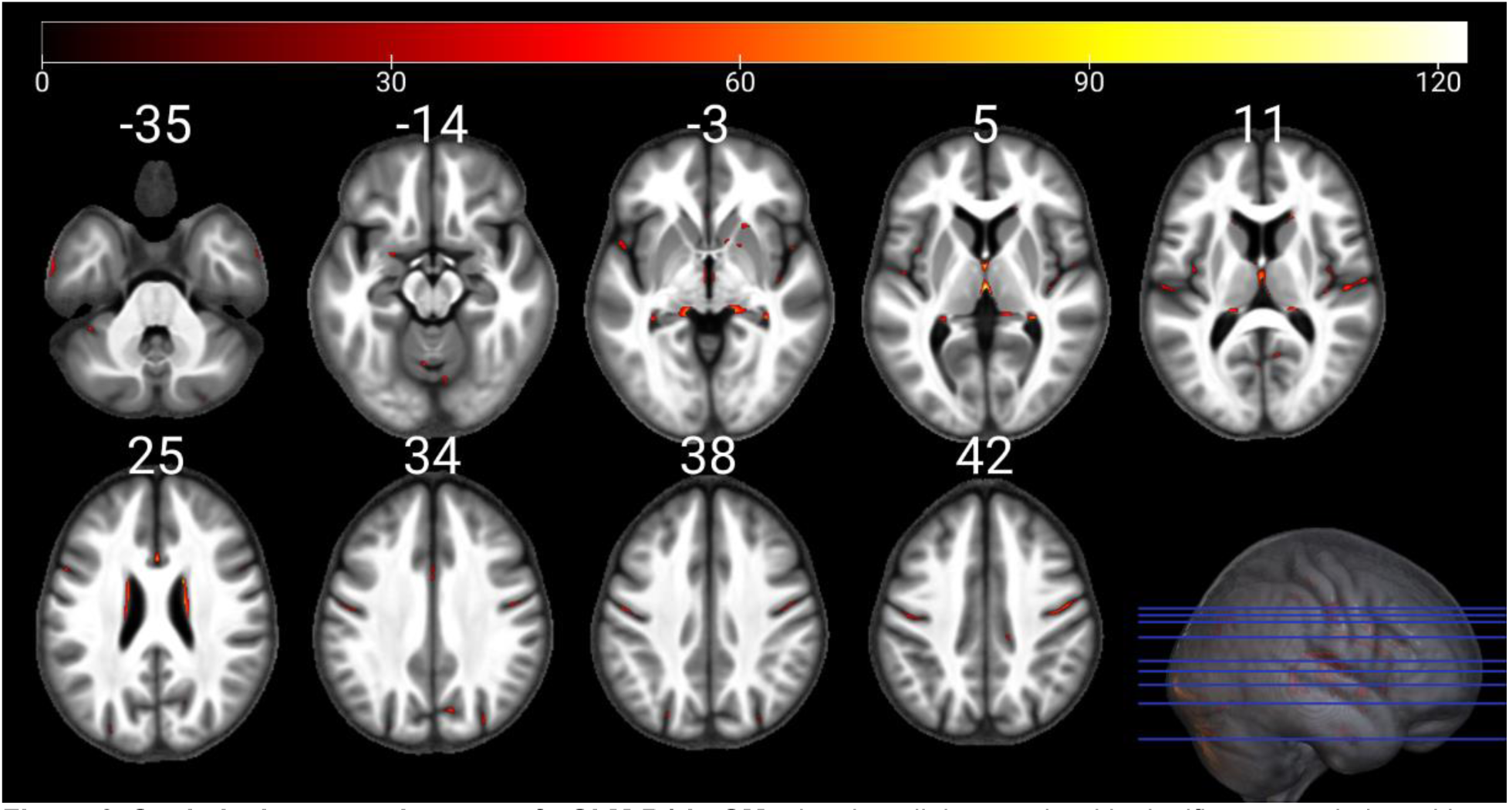
Statistical parametric maps of uGLM R1 in GM; showing all the voxels with significant correlation with age, as detected by uGLMs for R1 maps. The F-tests were thresholded at p<.0125 FWER corrected at voxel-level. The SPMs were overlayed on the mean MTsat map for the cohort in the MNI space. Abbreviation: GLM, general linear model; uGLM, univariate GLM; GM, gray matter; FWER, family-wise error rate; SPM, statistical parametric map.

**Figure 4.**
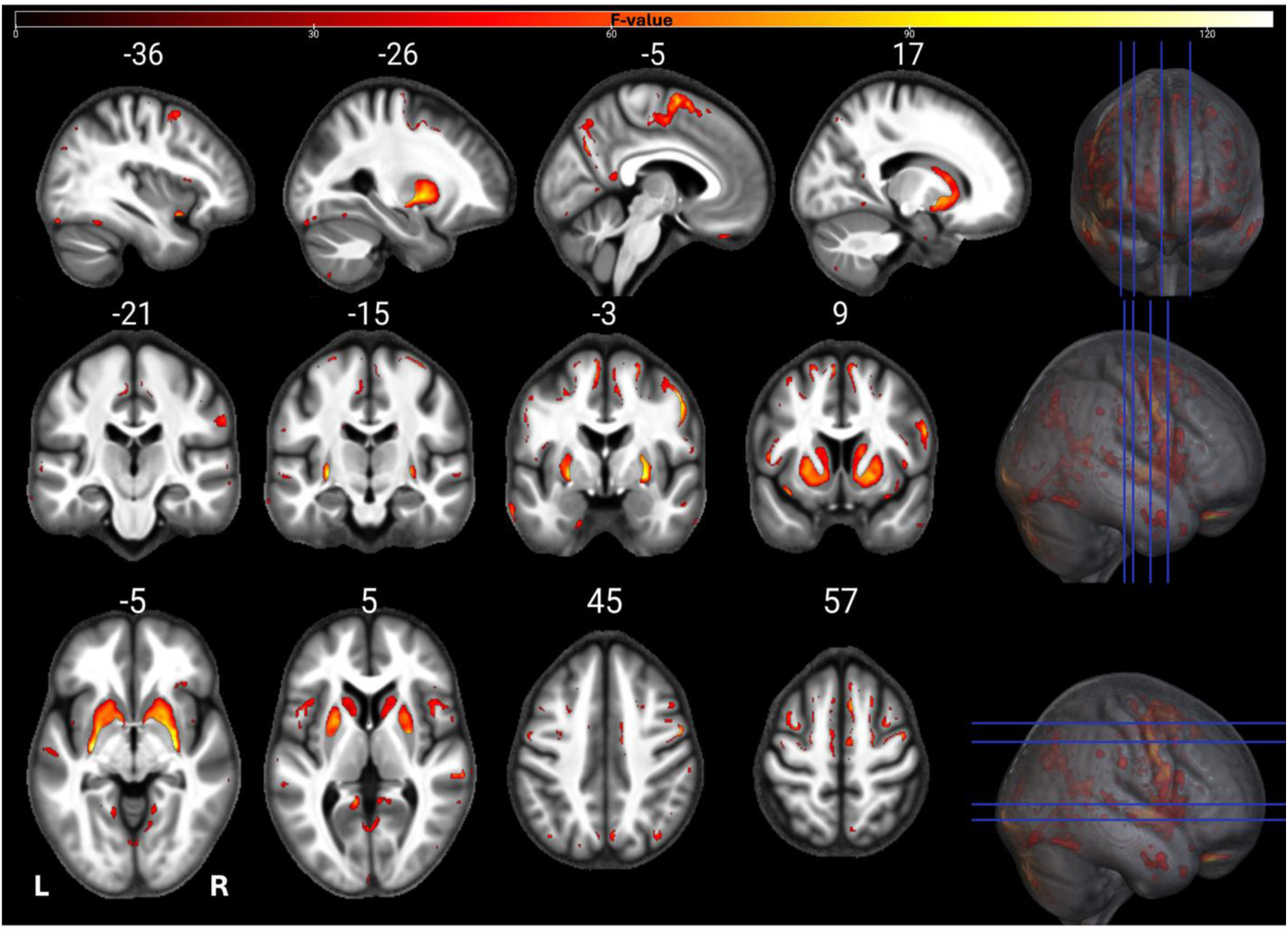
Statistical parametric maps of uGLM R2* in GM; showing all the voxels with significant correlation with age, as detected by uGLMs for R1 maps. The F-tests were thresholded at p<.0125 FWER corrected at voxel-level. The SPMs were overlayed on the mean MTsat map for the cohort in the MNI space. Abbreviation: GLM, general linear model; uGLM, univariate GLM; GM, gray matter; FWER, family-wise error rate; SPM, statistical parametric map.

**Figure 5.**
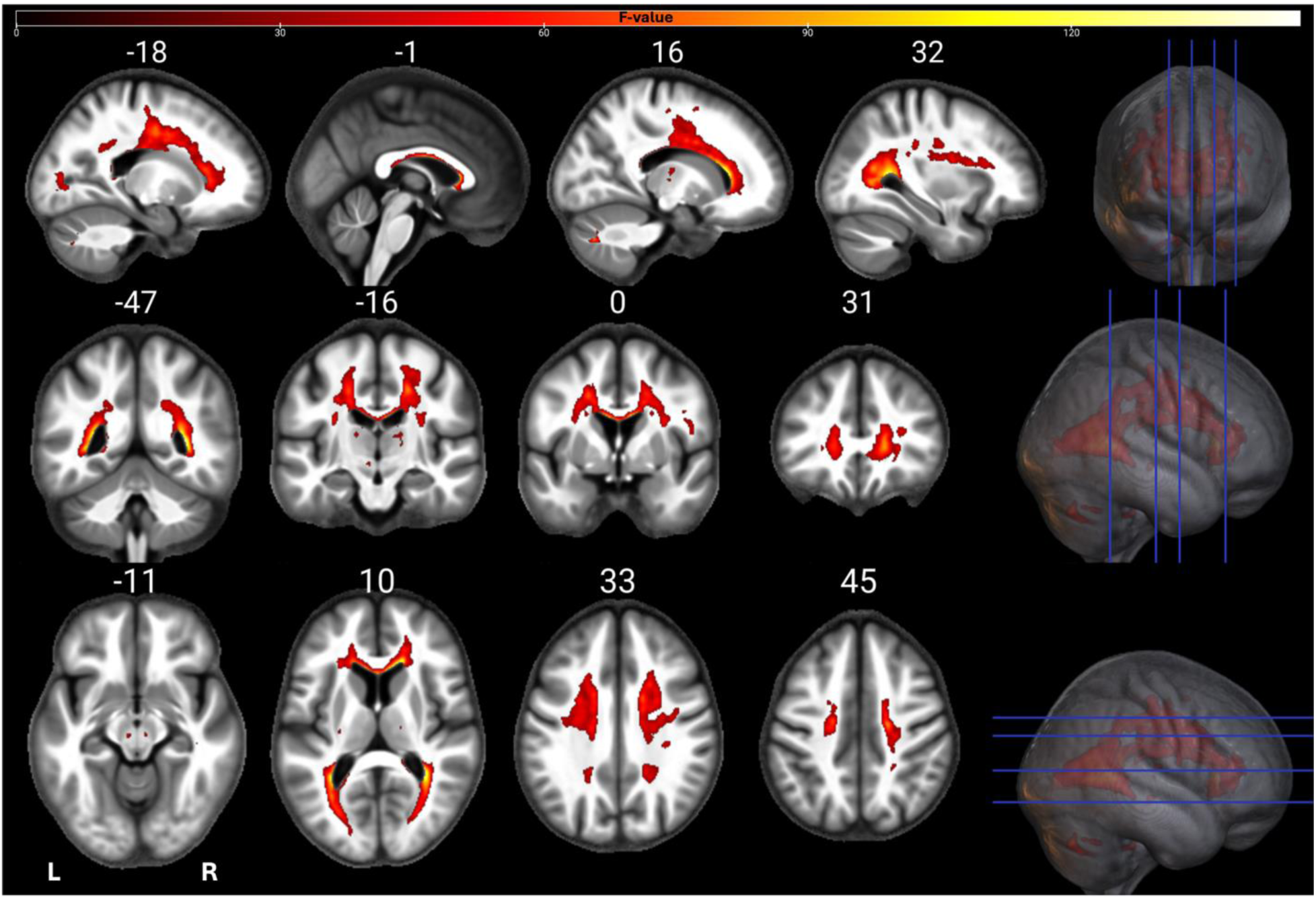
Statistical parametric maps of MTsat uGLM in WM; showing all the voxels with significant correlation with age, as detected by uGLMs for MTsat maps. The F-tests were thresholded at p<.0125 FWER corrected at voxel-level. The SPMs were overlayed on the mean MTsat map for the cohort in the MNI space. Abbreviation: GLM, general linear model, uGLM, univariate GLM, mGLM, multivariate GLM, WM, white matter, FWER, family-wise error rate, SPM, statistical parametric map.

**Figure 6.**
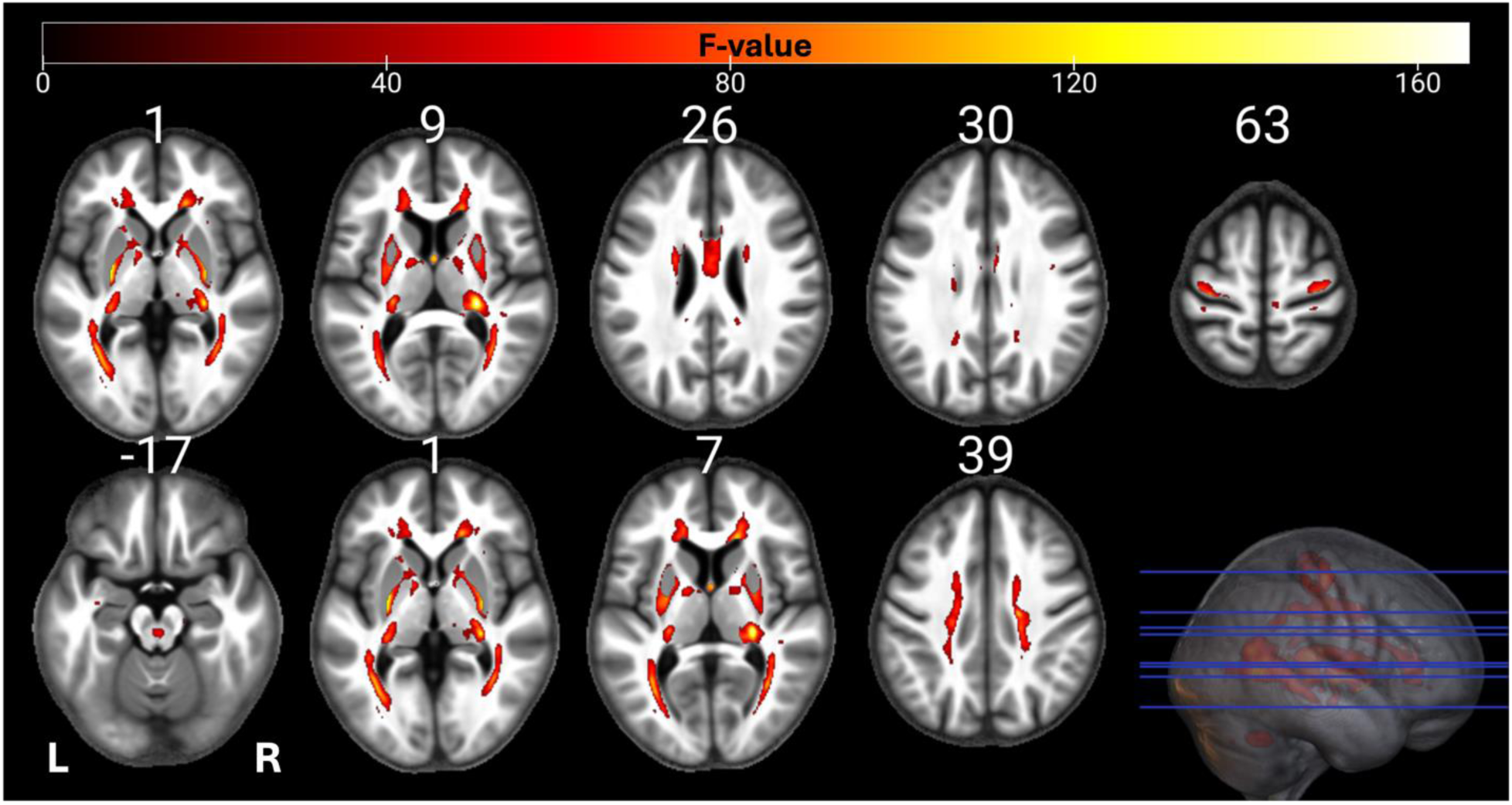
Statistical parametric maps of PD uGLM in WM; showing all the voxels with significant correlation with age, as detected by uGLMs for PD maps. The F-tests were thresholded at p<.0125 FWER corrected at voxel-level. The SPMs were overlayed on the mean MTsat map for the cohort in the MNI space. Abbreviation: GLM, general linear model, uGLM, univariate GLM, mGLM, multivariate GLM, WM, white matter, FWER, family-wise error rate, SPM, statistical parametric map.

**Figure 7.**
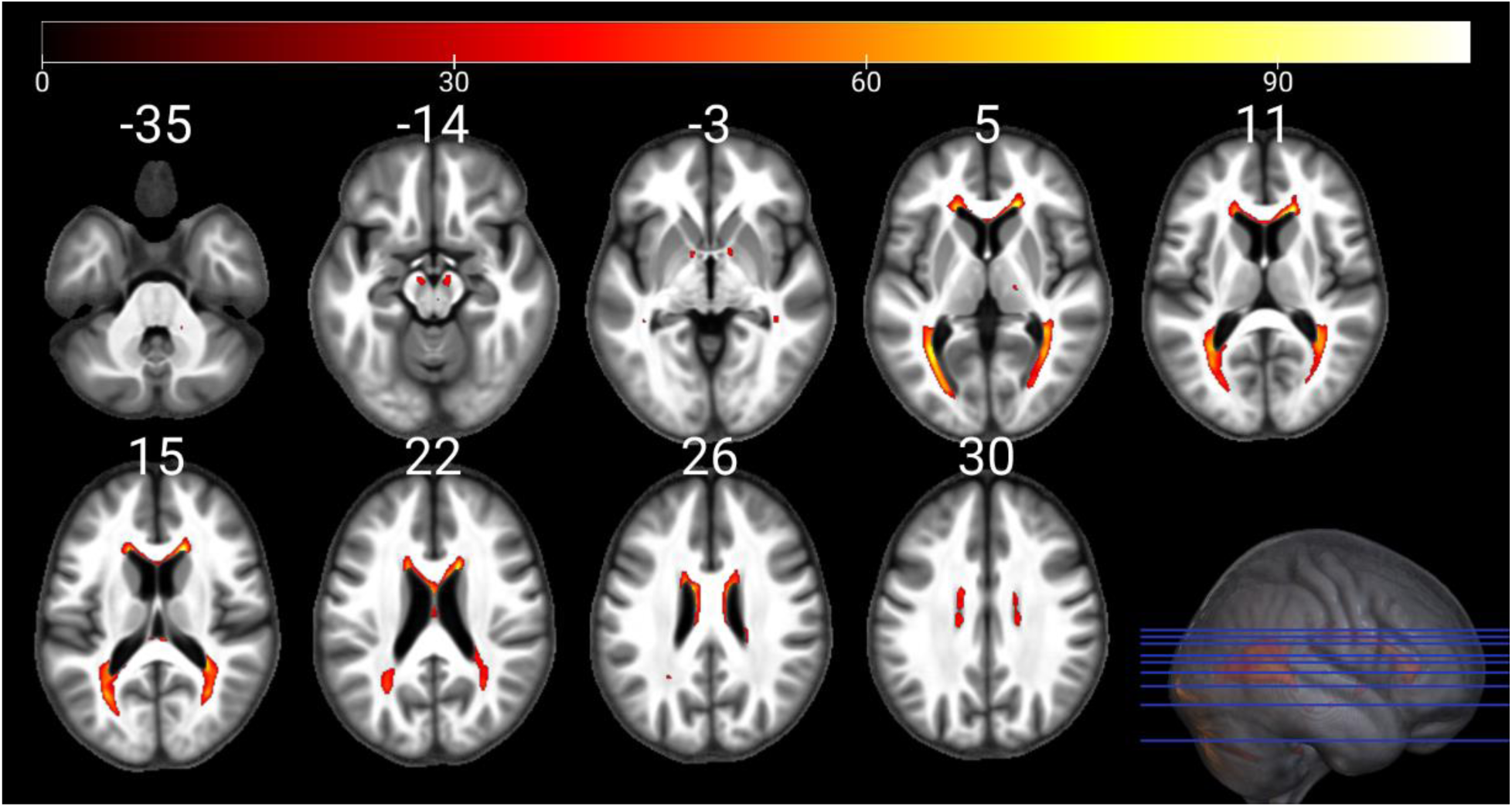
Statistical parametric maps of R1 uGLM in WM; showing all the voxels with significant correlation with age, as detected by uGLMs for R1 maps. The F-tests were thresholded at p<.0125 FWER corrected at voxel-level. The SPMs were overlayed on the mean MTsat map for the cohort in the MNI space. Abbreviation: GLM, general linear model, uGLM, univariate GLM, mGLM, multivariate GLM, WM, white matter, FWER, family-wise error rate, SPM, statistical parametric map.

**Figure 8.**
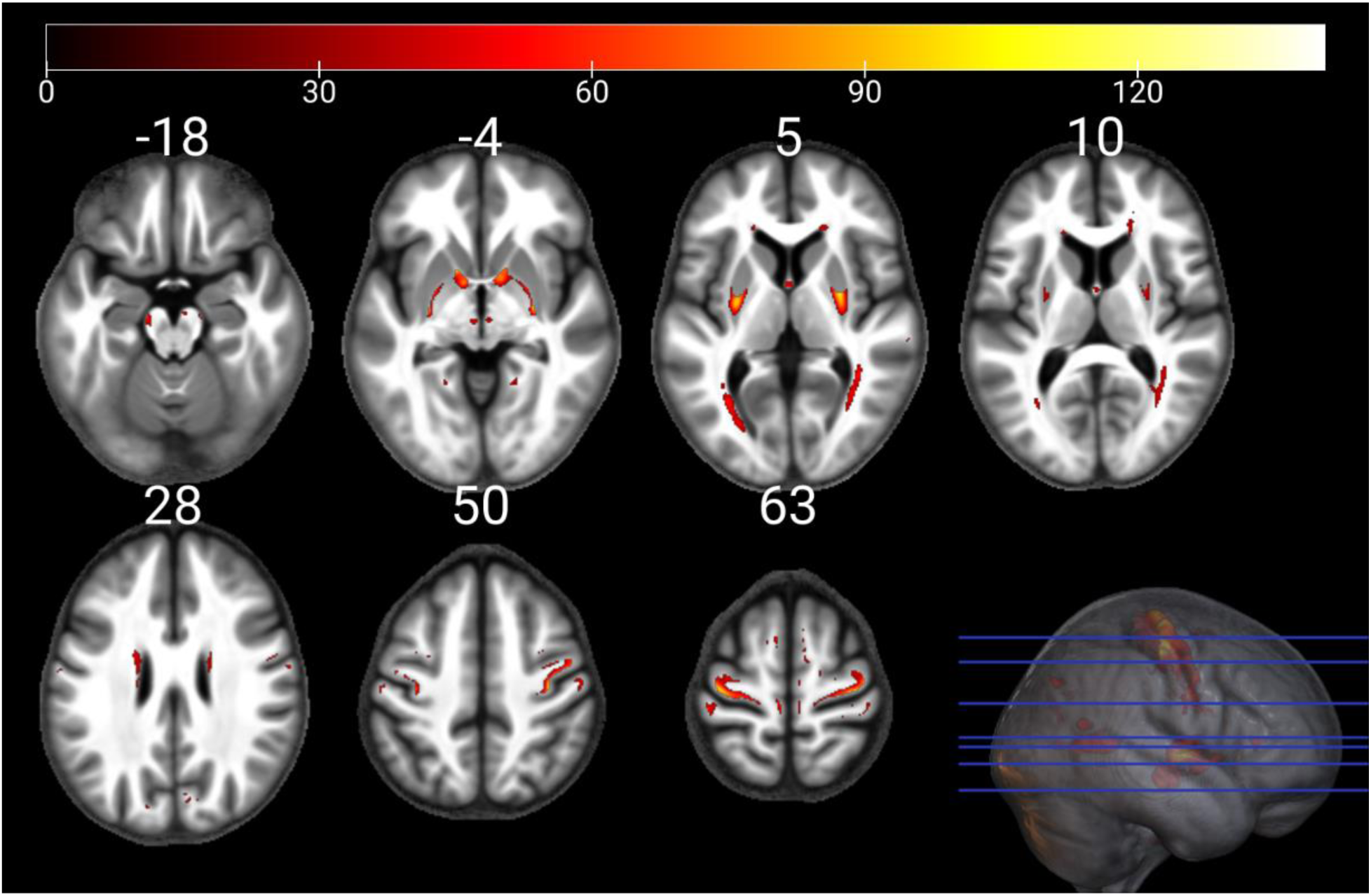
Statistical parametric maps of R2* uGLM in WM; showing all the voxels with significant correlation with age, as detected by uGLMs for R2* maps. The F-tests were thresholded at p<.0125 FWER corrected at voxel-level. The SPMs were overlayed on the mean MTsat map for the cohort in the MNI space. Abbreviation: GLM, general linear model, uGLM, univariate GLM, mGLM, multivariate GLM, WM, white matter, FWER, family-wise error rate, SPM, statistical parametric map.

**Figure 9.**
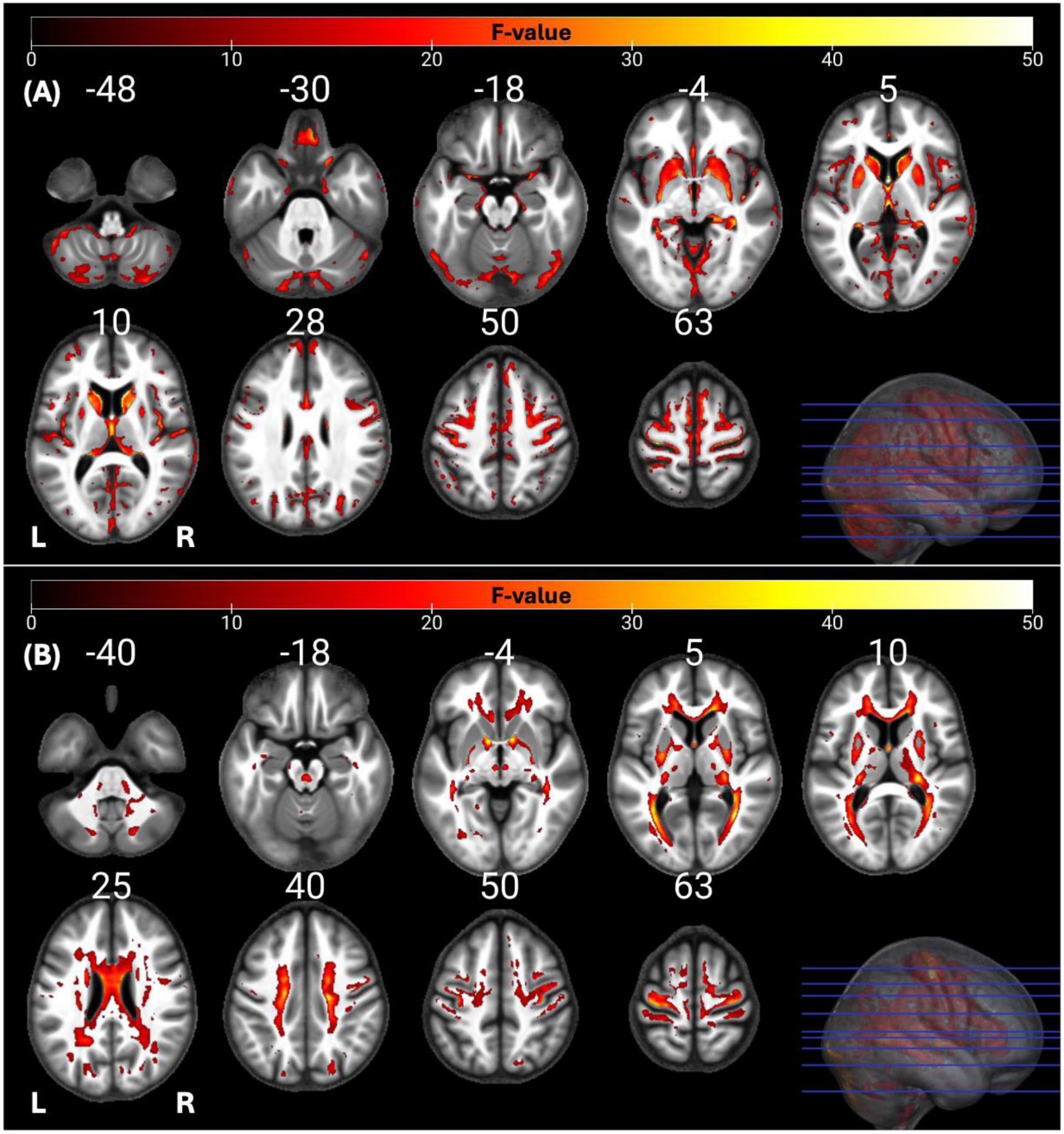
Statistical parametric maps of mGLMs in GM and WM; showing all the voxels with significant correlation with age, as detected by the multivariate model. The F-tests were thresholded at p<.05 FWER corrected at voxel-level. The SPMs were overlayed on the mean MTsat map for the cohort in the MNI space. Abbreviation: GLM, general linear model; mGLM, multivariate GLM; WM, white matter, FWER, family-wise error rate, SPM, statistical parametric map.

Table 1 provides a summary of the F-tests results (thresholded at voxel-level p<.05 FWER) from the univariate general linear models (uGLMs), union of these (UuGLMs) and mGLM for all maps within the two tissue classes. Additionally, Table 2 presents the summary of the uGLM F-tests results with Bonferroni-adjusted threshold, plus their union (UuGLMs). Comparing the spatial extent of clusters of significant voxels between uGLMs, UuGLMs and mGLM in Tables 1 and 2 reveals that the multivariate model (mGLM) identifies a larger number of significant voxels compared to each individual uGLMs, as well as their union (UuGLMs) with both p<.05 FWER and p<.0125 Bonferroni corrections. When thresholding the uGLMs at p<.05 FWER, the number of clusters observed with the mGLM is smaller than within the UuGLMs; though the mGLM clusters reach larger spatial extent than those of the UuGLMs. When thresholding the uGLMs at p<.0125 FWER, the mGLM results in more numerous and larger clusters then with the UuGLMs. To illustrate the voxels related to ageing at a threshold of p<.0125 and p<.05 in at least one of the (semi-)quantitative maps, the union of all thresholded statistical parametric maps derived from uGLMs in GM, i.e. UuGLMs, is depicted in **Figure *10***. In **Figure *11***, we then show in red the voxels that are uniquely detected by the mGLM, and in green, those uniquely detected by any of the uGLMs. Overall comparing mGLM vs. UuGLMs results, at both p_FWER_ <.05 FWER and p_FWER_<.0125 lead to Cohen’s Kappa values of .6599 and .6663 respectively in the grey matter and of .7199 and.7292 respectively in the white matter, which all can be interpreted as a “good” agreement (Landis and Koch, 1977).

**Figure 10.**
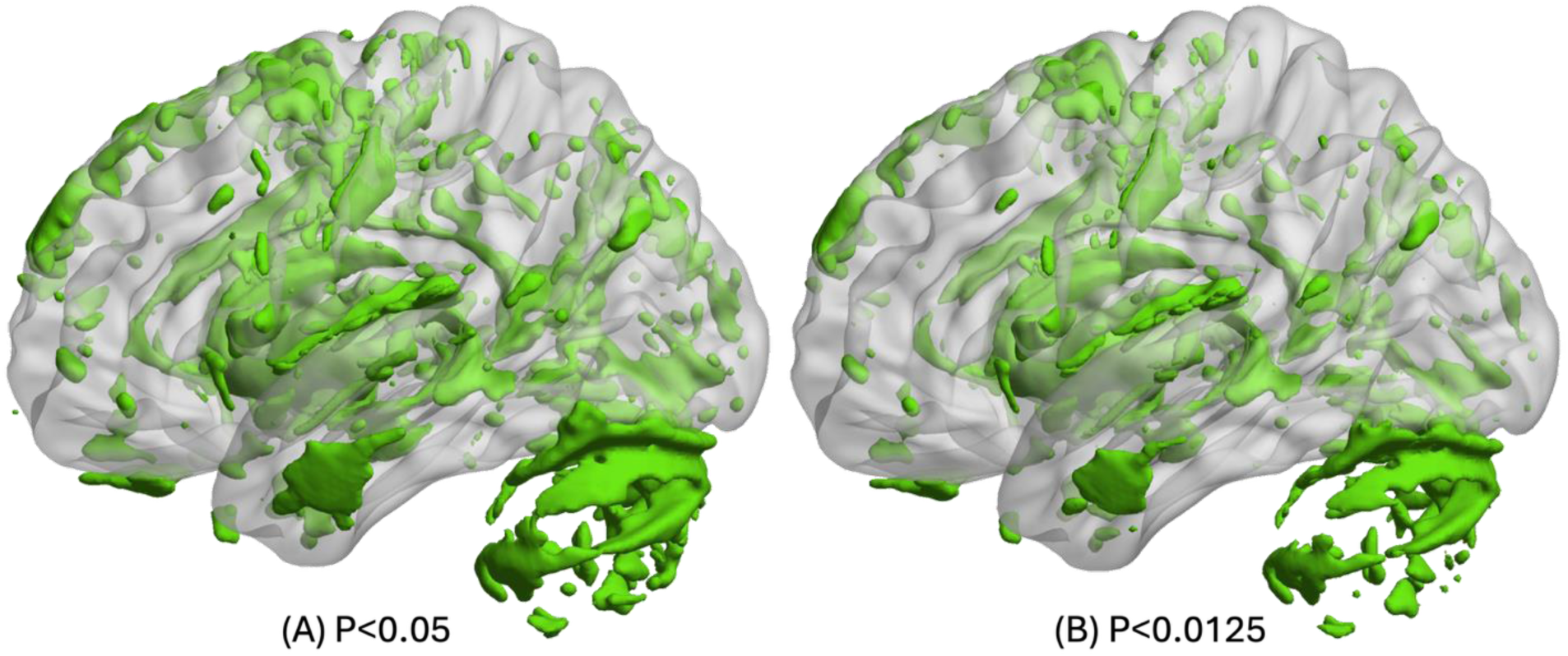
The union of all uGLM statistical parametric masks in GM. On the left side, the F-tests for the uGLMs were thresholded at at p<.05 FWER corrected at voxel-level. On the right side, the F-tests for the uGLMs were thresholded at P<.0125 after correction for FWER at voxel-level. The masks were overlayed on the mean MTsat map for the cohort in the MNI space. The right side of the brain is illustrated here. Abbreviation: GLM, general linear model; uGLM, univariate GLM; GM, gray matter; FWER, family-wise error rate.

**Figure 11.**
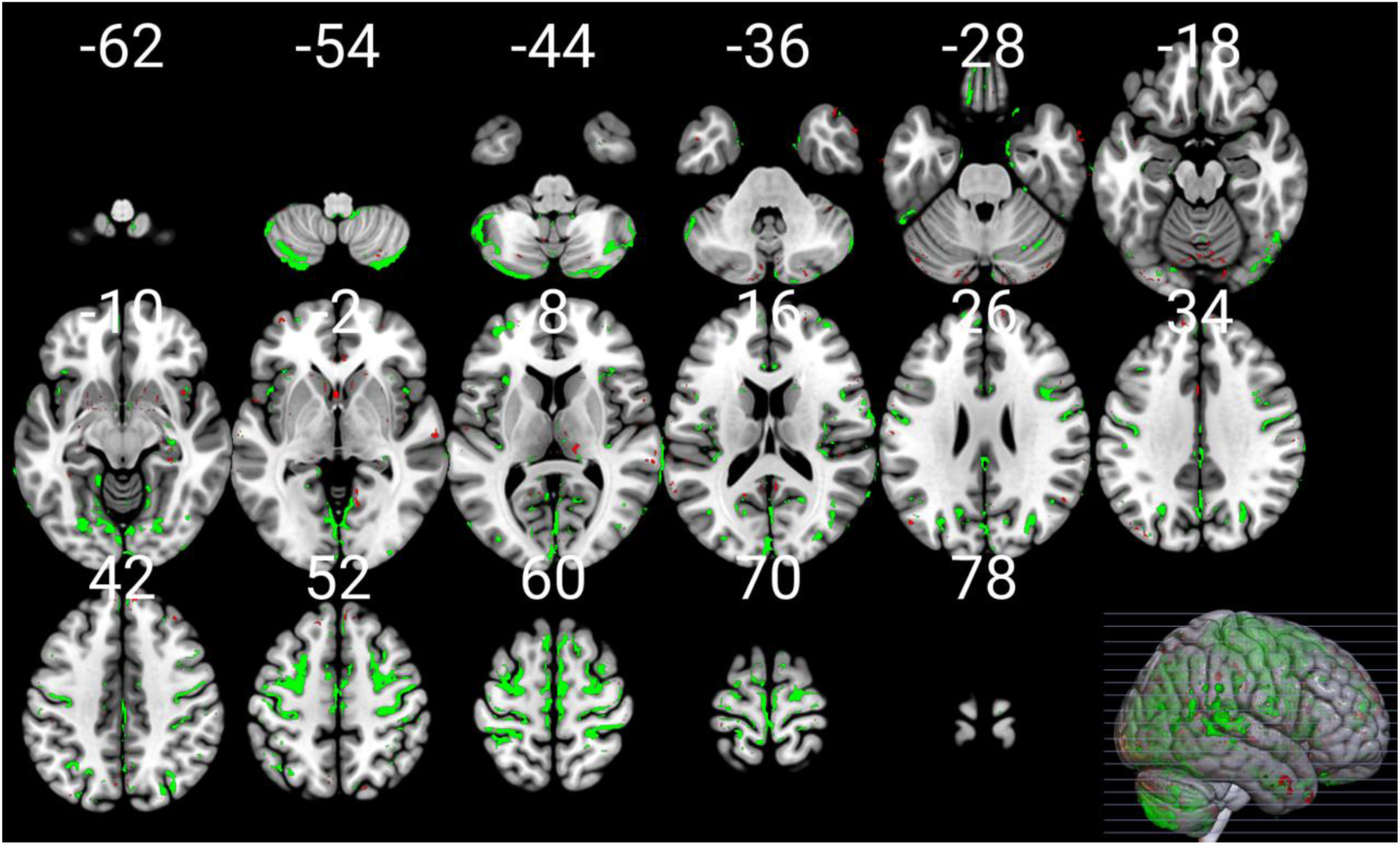
mGLM vs multiple uGLMs in GM. This figure shows the voxels detected either in the multivariate GLM model (thresholded at P<.05 FWER corrected at voxel-level in green), or in any of the univariate GLMs (thresholded at P<.0125 after correction for FWER at voxel-level in red). The mask is overlaid on the MNI152 template image in MRIcroGL. Abbreviation: GLM, general linear model; uGLM, univariate GLM; mGLM, multivariate GLM; GM, gray matter; FWER, family-wise error rate.

**Table 1.**
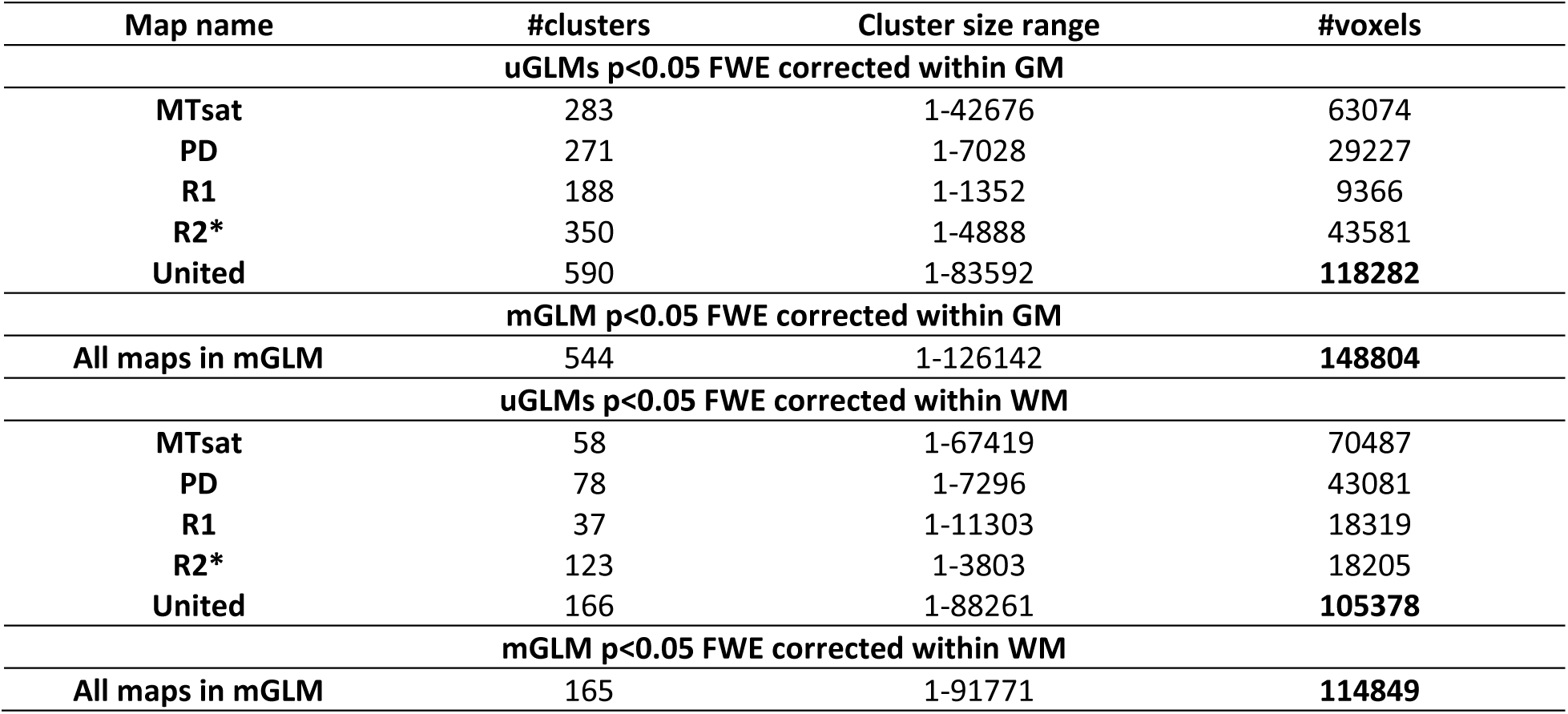
Summary statistics for significant voxels in uGLMs and mGLM. **“United” rows show the union of** significant voxels in SPMs for all modalities. Univariate GLMs were thresholded at p<0.05 FWER corrected per tissue class (GM or WM). Abbreviation: GLM, general linear model; uGLM, univariate GLM; mGLM, multivariate GLM; GM, gray matter; WM, white matter.

| Map name | #clusters | Cluster size range | #voxels |
| --- | --- | --- | --- |
| **uGLMs $p < 0.05$ FWE corrected within GM** |  |  |  |
| MTsat | 283 | 1-42676 | 63074 |
| PD | 271 | 1-7028 | 29227 |
| R1 | 188 | 1-1352 | 9366 |
| R2* | 350 | 1-4888 | 43581 |
| United | 590 | 1-83592 | **118282** |
| **mGLM $p < 0.05$ FWE corrected within GM** |  |  |  |
| All maps in mGLM | 544 | 1-126142 | **148804** |
| **uGLMs $p < 0.05$ FWE corrected within WM** |  |  |  |
| MTsat | 58 | 1-67419 | 70487 |
| PD | 78 | 1-7296 | 43081 |
| R1 | 37 | 1-11303 | 18319 |
| R2* | 123 | 1-3803 | 18205 |
| United | 166 | 1-88261 | **105378** |
| **mGLM $p < 0.05$ FWE corrected within WM** |  |  |  |
| All maps in mGLM | 165 | 1-91771 | **114849** |

**Table 2.** Summary statistics for significant voxels in uGLMs and mGLM. “United” rows show the union of significant voxels in SPMs for all modalities. Univariate GLMs were thresholded at p<0.0125=.05/4 FWER corrected, to account for the multiplicity of maps tested (4) per tissue class (GM or WM). The mGLM was thresholded at p<0.05 FWER corrected. Abbreviation: GLM, general linear model, uGLM, univariate GLM, mGLM, multivariate GLM, SPM, statistical parametric maps, GM, gray matter, WM, white matter.

| Map name | #clusters | Cluster size range | #voxels |
| --- | --- | --- | --- |
| **uGLMs $p < 0.0125$ FWE corrected within GM** |  |  |  |
| MTsat | 269 | 1-16578 | 50619 |
| PD | 220 | 1-6700 | 22596 |
| R1 | 150 | 1-677 | 6463 |
| R2* | 263 | 1-3632 | 32946 |
| United | 502 | 1-56154 | **92005** |
| **mGLM $p < 0.05$ FWE corrected within GM** |  |  |  |
| All maps in mGLM | 544 | 1-126142 | **148804** |
| **uGLMs $p < 0.0125$ FWE corrected within WM** |  |  |  |
| MTsat | 44 | 1-52986 | 55062 |
| PD | 73 | 1-6281 | 35295 |
| R1 | 27 | 1-8654 | 14366 |
| R2* | 100 | 1-3248 | 13650 |
| United | 138 | 1-60390 | **83428** |
| **mGLM $p < 0.05$ FWE corrected within WM** |  |  |  |
| All maps in mGLM | 165 | 1-91771 | **114849** |

Among the models used, only the mGLM detected an age effect in certain regions, including portions of the superior medial frontal lobe, supplementary motor area, paracentral lobule, middle and anterior cingulum, parts of the precuneus, cuneus, calcarine, lingual gyrus, cerebellum, hippocampus, and parahippocampal gyrus bilaterally, as well as the left fusiform gyrus.

### 3.2. Univariate and multivariate model evaluation

To assess the stability of the findings, the full analytical pipeline was repeated in two independent split-half samples (CV1 and CV2).

Compared to the full-sample analysis, all models (uGLMs, UuGLMs and mGLM) identified far fewer significant voxels (and clusters) in each split-half dataset, reflecting the reduced sample size and corresponding loss of statistical power. This reduction was more pronounced for the mGLM, which only showed larger voxel and cluster counts for the individual uGLMs but not for their union (UuGLMs). The match between the mGLM and UuGLMs maps of significant voxels was also reduced, compared to the full dataset with, Cohen’s Kappa values of about .52 on average (range from .4821 to .5382) for the grey matter and about .65 on average (range from .6237 to .6626) for the white matter, across both folds and uGLMs threshold (p<.05 and p>.0125 FWER). These can be interpreted as moderate and good agreement for the grey and white matter results respectively (Landis and Koch, 1977).

Importantly, ROI-based analyses demonstrated consistent effect sizes and directions of associations across CV1 and CV2, indicating that the underlying relationships captured by the uGLMs and mGLM were preserved, even when statistical power was reduced.

### 3.3. ROI analyses

We decided to perform Region of interest (ROI) analyses on the regions of interest listed in (Callaghan et al., 2014) to replicate previous findings. Moreover, we compared the canonical vectors withing the listed ROIs to investigate the contribution of each (semi-)quantitative map in those critical regions.

#### Median values

The median values of normalized, smoothed, and z-transformed R1, R2*, MTsat, and PD maps within the Putamen and Hippocampus ROIs with respect to age are illustrated in **Figure 12** and **Figure 13**. ROI-based regression analysis for the adjusted medians and age, within each cluster, is also provided, along with the respective linear age dependence observed from the uGLM analyses in the selected regions.

**Figure 12.**
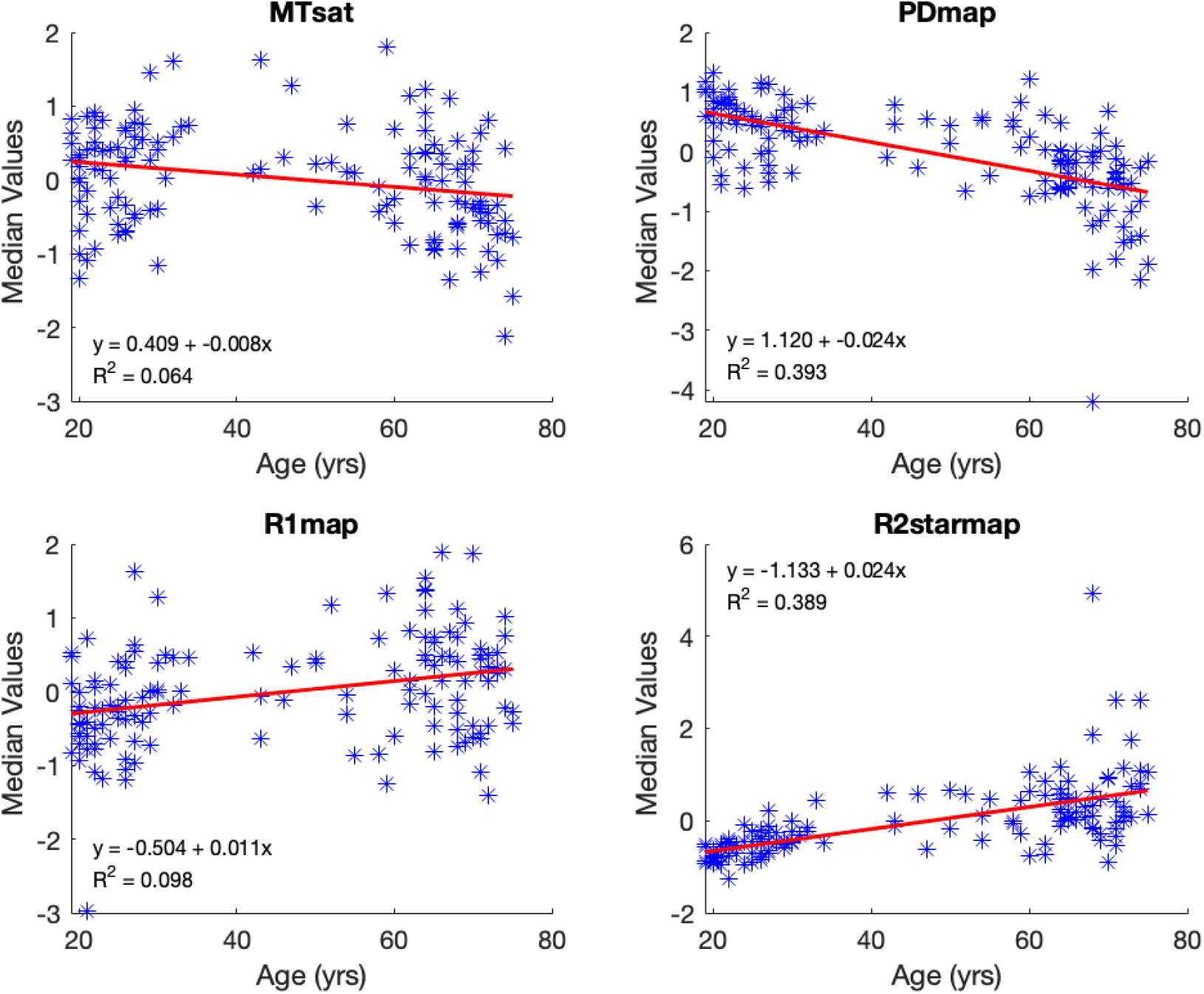
Median voxel values bilaterally in the Putamen. The red lines depict the linear model fit. These data are shown for illustration purposes only and were not used for any additional analyses.

**Figure 13.**
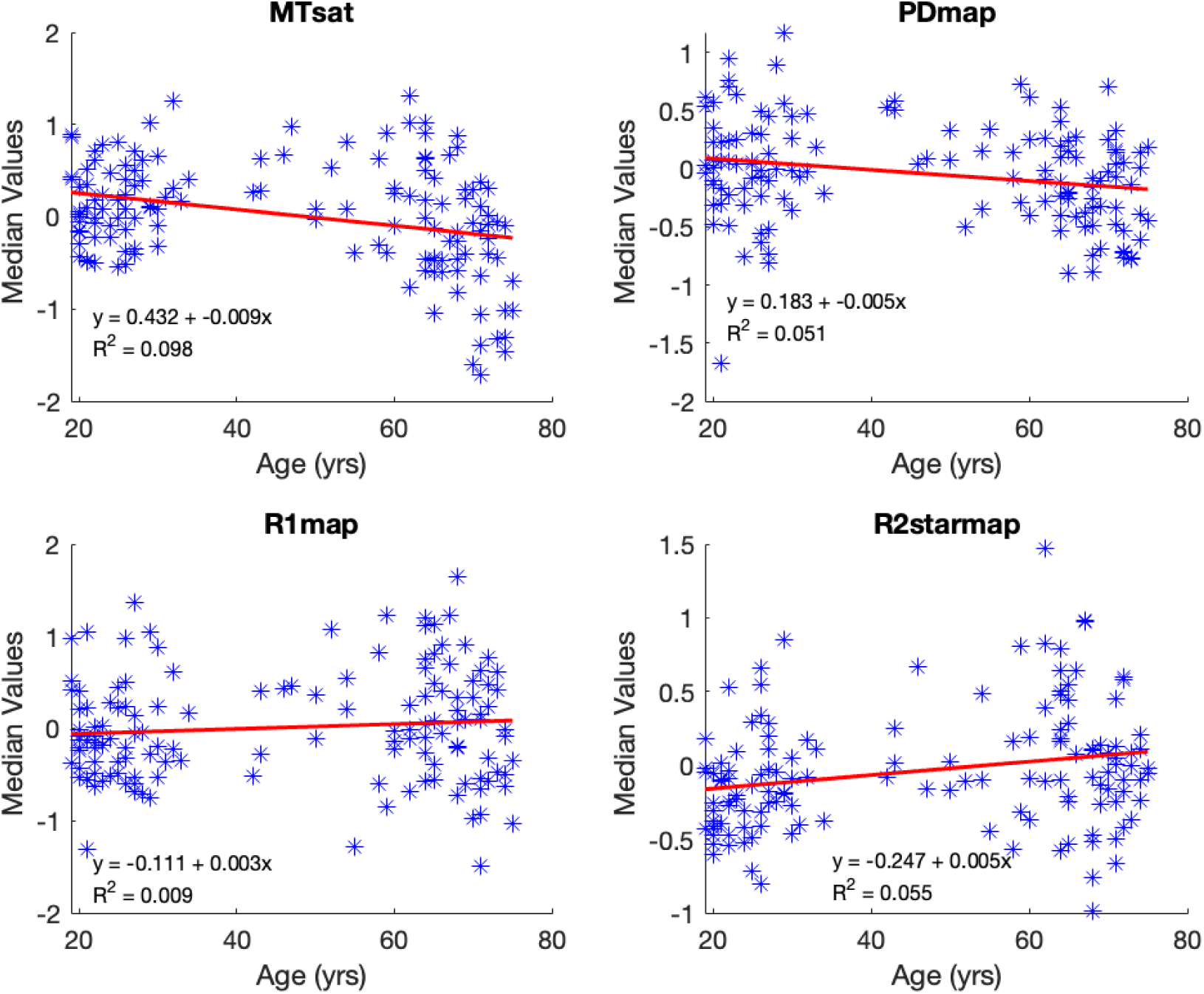
Median voxel values within the hippocampi. The red lines depict the linear model fit. These data are shown for illustration purposes only and were not used for any additional analyses.

From the ROI-based univariate regression analysis, we observe a bilateral decrease in the normalized MTsat and PD values in GM with respect to aging. While the PD and MTsat median values decrease with age in all regions of the brain, median R2* values show an increase in most regions of the brain except for the thalamus, where there is a significant negative age-related variation. These results are consistent with previously published results for the mean values in the thalamus for R2* (Taubert et al., 2020) but were not present in the uGLM results. R1 median signals show a weak positive correlation with age. The alteration in median values as a function of age concurs with the magnitude of the associated canonical vector. The bivariate correlation analysis indicates the strongest correlations for PD in most selected regions, see Table 3.

**Table 3.**
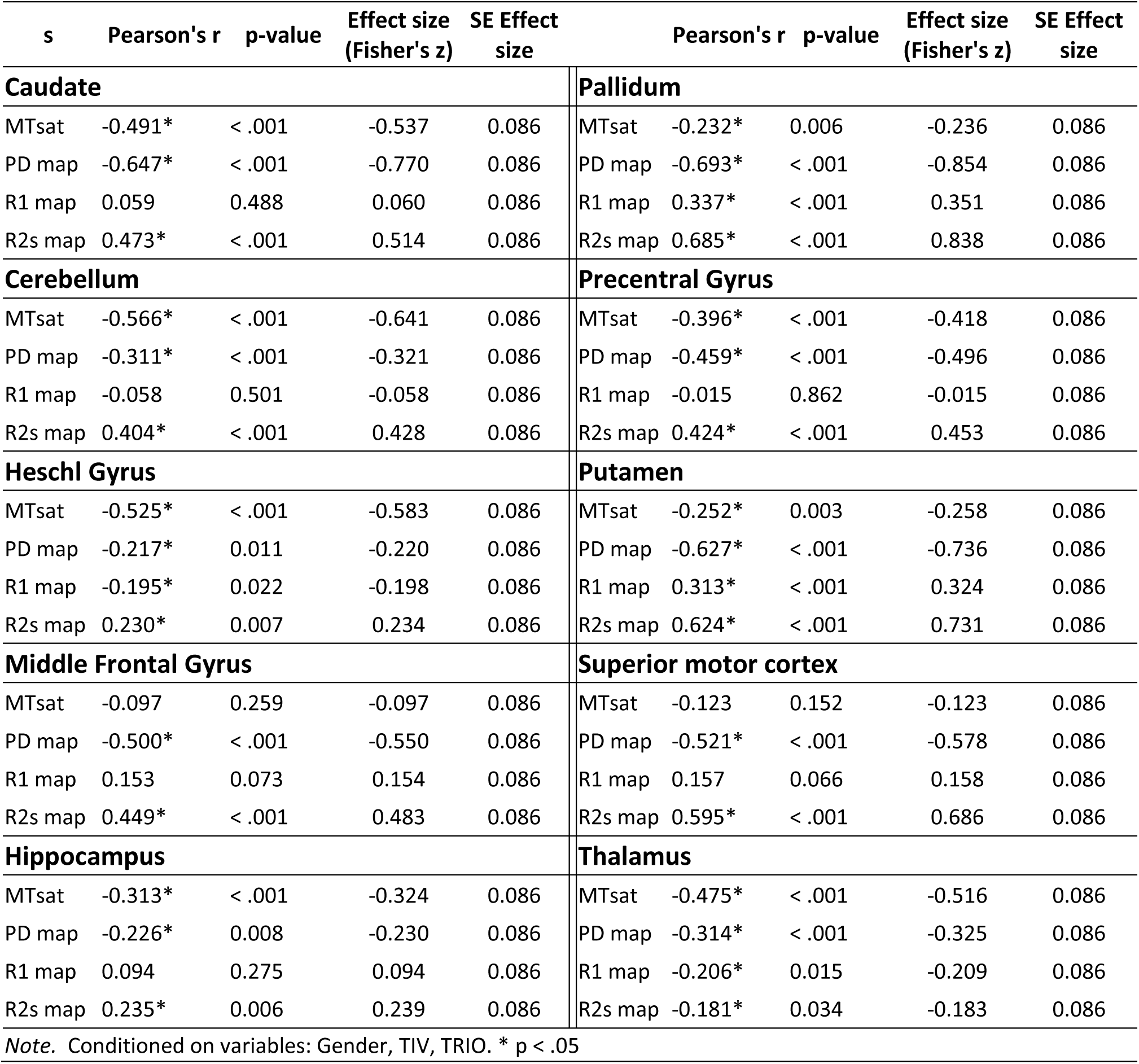
Pearson Partial Correlations on the median voxel values within different regions of interest.

#### Canonical vectors

To further investigate the impact of aging on myelination, iron content, and water concentration in various brain regions, (semi-)quantitative MR parameters were selected from bilateral regions including the putamen, thalamus, hippocampus, cerebellum, caudate, middle frontal gyrus, precentral gyrus, Heschl gyrus, supplementary motor area, caudate, and pallidum (Callaghan et al., 2014; Darnai et al., 2017; Tian et al., 2022; Wang et al., 2020).

For each ROI, the canonical vectors corresponding to each (semi-)quantitative map within the gray matter (GM) were estimated, as shown in the upper panel of **Figure 14**. It is important to note that the canonical vectors displayed in the lower part of **Figure 14** represent the contribution of each map in the peak voxel of the respective ROI and do not represent the contribution factor for the entire ROI. Additionally, the direction of these vectors cannot be interpreted as they are derived from F-tests; therefore, we have displayed the absolute weights. As illustrated, MTsat signals exhibit the highest contribution, while R1 and PD signals contribute the least in the mGLM as indicated by the canonical vectors.

**Figure 14.**
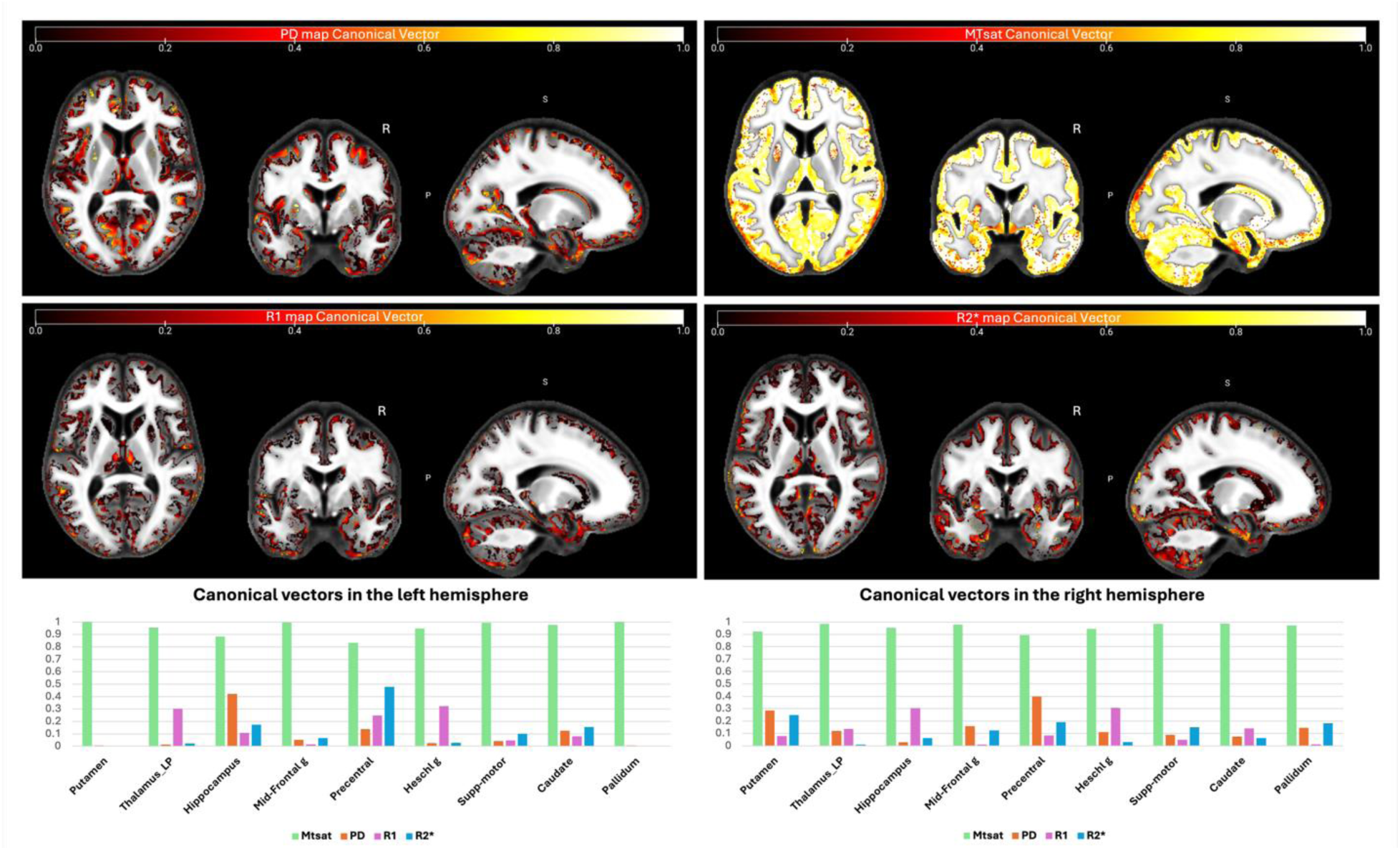
Canonical vectors for different modalities from the mGLM model, representing the contribution of each modality in each voxel. The color bars show the vector sizes [0,1]. The vectors correspond to the peak voxels at the selected ROIs. Detailed vector sizes are reported in Table sup-1 of the supplementary data. Abbreviation: mGLM, multivariate general linear model; ROI, region of interest.

## Discussion

The multivariate approach used in this study to investigate age-related changes in the microstructural tissue properties of the brain, incorporating image-derived (semi-)quantitative maps for myelin, iron, and free water content, enables the identification of regions that are influenced by the simultaneous occurrence of various parameter differences. The present findings should be interpreted as extending, rather than replacing, prior results obtained from this dataset. By applying a multivariate framework, we highlight the collective contribution of multiple imaging modalities to brain aging, offering a complementary view that builds on earlier univariate results.

The observed association with age in the quantitative MR parameters aligns with findings from *ex vivo* histologic studies and demonstrate a high specificity for tissue properties, including myelin content, iron content, and free water content. Using voxel-wise analysis with the multivariate GLM (mGLM), a bidirectional correlation between age and all the examined modalities was observed bilaterally in multiple brain regions. These regions encompassed the caudate nucleus, putamen, insula, cerebellum, lingual gyri, hippocampus, and olfactory bulb. Importantly, the multivariate approach demonstrated advantages over univariate analyses that focus on individual tissue parameters separately.

While examining individual tissue properties in isolation may provide insights into specific aspects of brain aging, the multivariate model revealed large clusters in the brain that could not be detected by analyzing each property individually. This indicates that the combined examination of multiple tissue properties enables the detection of additional regions associated with aging despite the presence of opposite changes in different properties. The multivariate patterns identified in this study likely reflect interacting age-related processes, including (toxic) iron accumulation, gradual myelin degradation, and changes in tissue water content. Iron-sensitive measure (R2*) highlight regionally heterogeneous patterns of iron accumulation, a process known to increase with age and to influence local tissue contrast, oxidative stress, and vulnerability to neurodegeneration. In parallel, myelin-sensitive MRI metric (MTsat) alterations further suggest gradual changes in tissue composition and water–myelin balance, aligning with evidence that myelin degradation and remodeling are early and continuous features of aging. Moreover, water content related variations (evident from PD maps), reflect the importance of structural connectivity as indexed by diffusion MRI studies in aging.

Importantly, the multivariate framework allows these tissue properties to be interpreted jointly, reflecting the fact that aging-related changes in microstructure, myelin, and iron are biologically interdependent rather than independent processes. As observed in Table 2, using the full dataset, mGLM showed improved sensitivity compared to the individual univariate GLMs (uGLMs) by detecting a larger number of significant voxels within clusters that cover the supplementary motor area, frontal cortex, hippocampus, amygdala, occipital cortex, and cerebellum bilaterally. This finding suggests that mGLM is a more effective and sensitive technique for detecting age-related differences in the brain, which should extend to other qMRI studies.

Using split-half datasets, for the cross-validation, sensitivity was reduced for all models, as expected but it appeared more so for the multivariate model, compared to the univariate models and the union of their results when the maps were thresholded at p<.05 FWER, which potentially points at a relatively increase risk of false positives within each individual map. Indeed, with the more conservative p<.0125 FWER threshold, the multivariate model displayed similar or slightly improved sensitivity, compared to univariate models (and the union of their thresholded maps).

In this study, the application of a multivariate model such as MANOVA proves advantageous due to the well-established correlations among brain tissue characteristics and the interrelated nature of (semi-)quantitative map values. By accounting for this inherent correlation, MANOVA effectively reduces potential biases in the results that could arise from using multiple ANOVAs. MANOVA’s ability to consider the complex relationships between multiple variables allows it to detect effects that might be smaller than those detectable by ANOVA, providing a more comprehensive understanding of the data.

Additionally, independent covariates can affect the relationship between the brain microstructure characteristics rather than influencing only a single variable; such patterns cannot be detected by the univariate models. Moreover, MANOVA offers a convenient means to manage the family-wise error rate when simultaneously analyzing multiple dependent variables, effectively reducing type-1 errors. In this study, where four different variables were examined concurrently, better results were achieved under the same p-value threshold compared to multiple univariate analyses on the same GM and WM maps. This underscores the robustness and appropriateness of a multivariate approach for understanding the intricate relationships within the dataset. Here, we utilized Bonferroni threshold for combining the p-values to build up the same power for univariate and multivariate techniques. However, there are other methods for combining p-values for detection of partial association such ad Fisher and weighted Fisher methods, which are more relevant in case of many comparisons (Yoon et al., 2021).

The canonical vectors shown in **Figure 14** indicate the relative contribution of each modality to the multivariate model at the peak voxels in the selected ROIs. As these vectors are derived from multivariate F-tests, their directionality cannot be interpreted, and only the magnitude of the weights is meaningful. Consequently, the canonical vectors should not be viewed as direct biological effect sizes or region-wide contribution measures, but rather as indicators of parameter sensitivity within the multivariate model.

Within this framework, MTsat exhibits the largest canonical weights across most ROIs, suggesting that myelin-sensitive signal components are a dominant contributor to age-related multivariate effects. PD also shows substantial contributions in several regions. In contrast, R2* and R1 exhibit smaller canonical weights, indicating lower sensitivity within the multivariate context, despite showing significant age associations in ROI-based univariate analyses.

Here are the region-specific interpretations of MRI change co-occurrences:

### Basal Ganglia (Caudate, Putamen, Pallidum)

Across basal ganglia nuclei, aging is characterized by a consistent decrease in MTsat and PD, alongside a robust increase in R2*. The strong negative correlations between age and PD and MTsat suggest reductions in tissue water–myelin balance and myelin-related signal components, while the positive age-related increase in R2* is consistent with iron accumulation, a well-established feature of subcortical aging. Notably, R1 shows region-dependent behavior, with positive age correlations in the putamen and pallidum but weaker or nonsignificant effects in the caudate, indicating heterogeneous sensitivity to microstructural and compositional changes within basal ganglia subregions. While univariate analyses emphasize the dominance of PD and R2* effects, the multivariate framework captures their joint contribution, highlighting the coupling between iron accumulation and myelin- and water-related alterations across basal ganglia structures.

### Sensorimotor Cortex (Precentral Gyrus, Superior Motor Cortex, Heschl’s Gyrus)

Sensorimotor regions show a more nuanced aging profile. PD and MTsat generally decrease with age, indicating subtle changes in tissue composition and myelin-related properties, while R2* increases are consistently observed, albeit with moderate effect sizes. R1 effects are weak or nonsignificant in most sensorimotor ROIs. This pattern aligns with evidence that primary sensory and motor cortices exhibit relative structural preservation with aging but still undergo microstructural and compositional changes. The multivariate framework reveals that these modest regional effects participate in broader aging-related tissue-property patterns that are not easily detected through univariate voxel-wise analysis alone.

### Cerebellum

The cerebellum demonstrates significant age-related decreases in MTsat and PD and a positive association between age and R2*. These findings suggest concurrent myelin-related changes and iron-sensitive alterations, consistent with emerging evidence that the cerebellum undergoes nontrivial microstructural and compositional aging despite relatively preserved volume. R1 shows no significant age dependence, further emphasizing the differential sensitivity of qMRI parameters. While cerebellar effects are spatially heterogeneous in univariate analyses, the multivariate approach captures their integration into a global aging pattern involving both cortical and subcortical regions.

Overall, these region-specific results demonstrate that aging-related changes in iron-sensitive (R2*), myelin-sensitive (MTsat), and water-sensitive (PD) metrics occur in coordinated yet regionally heterogeneous patterns. By integrating these parameters simultaneously, the multivariate framework reveals biologically meaningful tissue-property interactions that extend beyond what can be inferred from univariate analyses alone. Finally, the canonical vectors are best interpreted as highlighting which tissue-sensitive MRI properties most strongly drive the multivariate age effect, rather than as mechanistic or directional markers of specific biological processes.

### Limitations and conclusion

It is important to note that MANOVA differs from multiple ANOVA analyses, as it does not focus on the signal-to-noise ratio of independent variable effects on each dependent variable individually. Instead, it tests for effects of interest on a combination of outcome variables. For an assessment of the former, a return to univariate analyses is necessary. Aging involves not only microstructural changes in the brain but also macrostructural and functional alterations, such as changes in GM and WM volume and cognitive behavior changes. Therefore, there remains a need to investigate aging by considering different combinations of changes together. Furthermore, longitudinal studies are required to elucidate how normal aging deviates from pathological aging within a multivariate system.

The nonlinear relationship between MTsat values and age, as visible in **Figure 12** and **Figure 13**, could be explained by the fact that myelination is not limited to early development, but can also occur throughout adulthood, with the pattern of myelination depending on the hierarchy of connections between different brain regions (Peters, 2002; Snaidero and Simons, 2014; Timmler and Simons, 2019). Potential nonlinear age dependency is also seen in the R2* profile in some regions such as amygdala and putamen (**Figure 13**) which could be due to slower accumulation of iron in these regions. Hagiwara et al. showed the nonlinear behavior of different brain tissue properties (Hagiwara et al., 2021). However, within the scope of this study, we limited our examination to linear age-related variations, as in the original analysis of these data. Moreover, the sampling of ages is too non-uniformly distributed, with the majority being young (<30 y.o.) and elderly (>50 y.o.) subjects to properly capture non-linear effects of aging.

In summary, the findings of this study underscore the importance of employing advanced statistical models like the mGLM to detect subtle microstructural changes associated with aging, when using multiple maps or multimodal data. The results highlight the significance of ROI analyses in identifying specific brain regions affected by aging and their relationships with different modalities. This study provides valuable insights into the neural mechanisms underlying age-related differences in brain structure, offering a multivariate framework for identifying distributed brain tissue patterns associated with normal aging, which may inform future precision-oriented studies of neurodegenerative risk.

## Supporting information

Supplementary methods and results

## Data Availability

The processed data used in this study are available online as well as the code to reproduce the results.

https://doi.org/10.18112/openneuro.ds005851.v1.0.0

https://github.com/CyclotronResearchCentre/qMRI_MSPM_AgeFx

## Data & Code Availability

The data supporting this study is available on Open Neuro: https://doi.org/10.18112/openneuro.ds005851.v1.0.0 . Moreover, the scripts used to generate the results presented in this study are provided on GitHub (https://github.com/CyclotronResearchCentre/qMRI_MSPM_AgeFx) as part of the replication package.

## Acknowledgments

The authors would like to thank E. Anderson, M. Cappelletti, R. Chowdhury, J. Diedirchsen, T.H.B. Fitzgerald, and P. Smittenaar, who took part in data acquisition as part of multiple cognitive neuroimaging studies performed at the WCHN.

## Supplementary

Figures S1-S7, Tables S1-S7

