## Supplementary methods and results for "Multivariate age-related variations in quantitative MRI maps: Widespread age-related differences revisited"

Christophe Phillips

GIGA – CRC Human Imaging, University of Liège

Cyclotron Research Centre, B30,

8 Allée du Six Août, 4000 Liège, Belgium

### SUPPLEMENTARY MATERIAL

### 1 Data acquisition parameters

As mentioned in (Callagahn et al. 2014), participants were examined on two 3T whole body MR systems (Magnetom TIM Trio, Siemens Healthcare, Erlangen, Germany, 69 participants per scanner) each equipped with a standard 32 channel head coil for receive and RF body coil for transmission. The data were acquired as part of several cognitive neuroimaging studies at the Wellcome Trust Centre for Neuroimaging with approval from the local ethics committee.

A whole-brain quantitative MPM protocol was used. This consists of 3 spoiled multi-echo 3D fast low angle shot (FLASH) acquisitions with 1 mm isotropic resolution and 2 additional calibration sequences to correct for inhomogeneities in the RF transmit field (Lutti et al., 2010, 2012; Weiskopf et al., 2013). The FLASH volumes were acquired with predominantly proton density (PD), T1 or MT weighting, determined by the repetition time, and flip angle ($\alpha$) (repetition time and flip angle were for the PD- and MT-weighted acquisitions: 23.7 ms/6°; and for the T1-weighted acquisition: 18.7 ms/20°). In the case of the MT-weighted acquisition, a Gaussian RF pulse with 4 ms duration and 220° nominal flip angle was applied 2 kHz off-resonance before nonselective excitation. Gradient echoes were acquired with alternating readout gradient polarity at 6 equidistant echo times between 2.2 ms and 14.7 ms. Two additional echoes were acquired for the PD-weighted acquisition at 17.2 ms and 19.7 ms. A high readout bandwidth of 425 Hz/pixel was used to reduce off-resonance artefacts (Helms and Dechent, 2009). To speed up data acquisition, parallel imaging with a speed up factor of 2 was used in the phase-encoded direction (anterior-posterior) using the generalized auto-calibrating partially parallel acquisition algorithm. A partial Fourier acquisition (6/8 sampling factor) was used in the partition direction (left-right). The total scanning time of the MPM protocol was approximately 25 minutes.

The total sum-of-square-and-cross-product (SSCP) matrix of the model is $SSCP_{Total} = Y'Y - N\bar{y}\bar{y}'$ and can be partitioned into the regression and residual SSCP:

$$SSCP_{Total} = SSCP_{Regression} + SSCP_{Residual} = (\hat{y}'\hat{y} - N\bar{y}\bar{y}') + E'E$$

**Hypothesis testing**

Hypothesis testing in mGLM, relies on testing the linear contrast $CBL = 0$. This extension of the univariate scheme combines standard hypotheses on the rows of matrix $B$, i.e. on the regressor, represented by matrix $C$, with hypotheses on the columns of $B$, i.e. on the different modalities, represented by matrix $L$. In the context of multivariate ANOVA (MANOVA) models, contrasts of main effects and interactions are formulated by setting $L = I_t$, the $t \times t$ identity matrix, as the dependent variables in multimodal neuroimaging applications are not assumed to be directly proportional. This method is the most suitable for conducting hypothesis testing on multimodal neuroimaging applications.

The SSCP matrices are defined as:

$$SSCP_{hypo} = \left(C\hat{B}L^T\right)^T \left(C(X^T X)^{-1} C^T\right)\left(C\hat{B}L^T\right)$$

And an SSCP matrix associated with the residuals:

$$SSCP_{residual} = L(\hat{E}^T E)L^T$$

**Test statistics in mGLM**

The global significance of the GLM, i.e. $H_0: B = 0$, one can simply be estimated as:

$$SSCP_{Regression} SSCP_{Residual}{}^{-1} = \frac{(\hat{y}'\hat{y} - N\bar{y}\bar{y}')}{E'E}$$

Then we can calculate the eigenvalues of the resulting matrix by solving $det\left(\frac{(\hat{y}'\hat{y} - N\bar{y}\bar{y}')}{E'E} - \lambda I_m\right) = 0$ to get the $m = 4$ eigenvalues.

In hypothesis testing using the multivariate GLM, there are four standard test statistics available, which can be constructed based on the calculation of the SSCP matrices: Pillai's trace (Pillai, 1955), Wilks' lambda (Wilks, 1932), Hotelling-Lawley trace (Hotelling, 1951), and Roy's largest root (Roy, 1945). The Wilks Lambda statistic can be calculated based on the calculated m eigenvalues as $\Lambda = \prod_{i=1}^{m} \frac{1}{1+\lambda_i}$. This has an approximate F distribution with degrees of freedoms $a$ and $b$:

$$F(a, b) = \frac{1 - \Lambda^{\frac{1}{t}}}{\Lambda^{\frac{1}{t}}} \cdot \frac{b}{a}$$

Where $a = lq$ and $b = rt - 2u$.

$$l = rank(L)$$

$$q = rank(C)$$

$$u = \frac{a - 2}{4}$$

$r = N - q - \frac{a+1}{2}$, N = sample size

$$t = \begin{cases} \sqrt{\dfrac{l^2 q^2 - 4}{l^2 + q^2 - 5}} & , l^2 + q^2 - 5 > 0 \\ 1 & , l^2 + q^2 - 5 \leq 0 \end{cases}$$

If the minimum value between $l = 4$ and $q = 1$ is $\leq 2$ , the distribution is exactly F.

The hypothesis $H_0: CBL = 0$ can be tested to assess the different potential co-occurrence of change between the modalities. To test the joint effect on the 4 quantitative maps in a specific tissue type, $L_{4\times4}$ is defined as an identity matrix corresponding to the number of dependent variables (semi-quantitative maps). As explained before, each column of $L$ will perform a univariate analysis on each column of $B$. Here, $C$ was defined as $[0\ 1\ 0\ 0\ 0]$ to only see the correlation between age and maps. In this case, $\Lambda$ has an exact F distribution with $a = 4$ and $b = 130$ degrees of freedom.

These matrices are generalizations of the numerator and denominator sums-of-squares from the univariate GLM hypothesis-testing approach. When $L$ is an identity matrix, the main diagonal of $SSCP_{hyppo}$ contains the sums of squares for the hypothesis in C as applied to the estimated parameters for each dependent variable separately. And the $SSCP_{residual}$ matrix is an unscaled form of the estimated covariance matrix $\hat{\Sigma}$.

Construction of the test statistics rely on some linear combination of $m$ eigenvalues $(\lambda_1, \dots, \lambda_q)$ of $SSCP_{residual}^{-1} SSCP_{hyppo}$. Here, we will only focus on Wilks' lambda test statistics. It quantifies the proportion of variance not accounted for by the hypothesis compared to the total variance in the data.

#### 2.3 Canonical Correlation Analysis

Canonical vectors are calculated under the assumption that matrix $L$ involves multiple dependent variables $(l > 1)$, to extract the contribution of each dependent variable to the test statistics $\Lambda$. This contribution corresponds to the eigenvectors of the eigen decomposition of $SSCP_{residuals}^{-1} SSCP_{hyppo}$ (Tabachnick and Fidell, 2007). Once linear combinations of canonical variates for the dependent $(U_1 = a_1y_1 + a_2y_2 + a_3y_3 + a_4y_4)$ and independent $(V1 = b_1x_1 + b_2x_2 + b_3x_3 + b_4x_4 + b_5x_5)$ variables are formed, the first canonical correlation is calculated by solving $det\left(\frac{(\hat{y}'\hat{y} - N\bar{y}\bar{y}')}{E'E} - \lambda I_m\right) = 0$ for $\lambda_1$. After finding the first pair of canonical variates, the process continues. The analysis finds a second pair of variates ($U_2$ and $V_2$) that are also maximally correlated with each other, but with the added constraint that they are uncorrelated with the first pair of variates. This process continues until a certain number of canonical variate pairs are extracted. The number of possible pairs is equal to the number of variables in the smaller of the two sets (in our case, 4).

### 3 Results

#### 3.1 Univariate GLMs

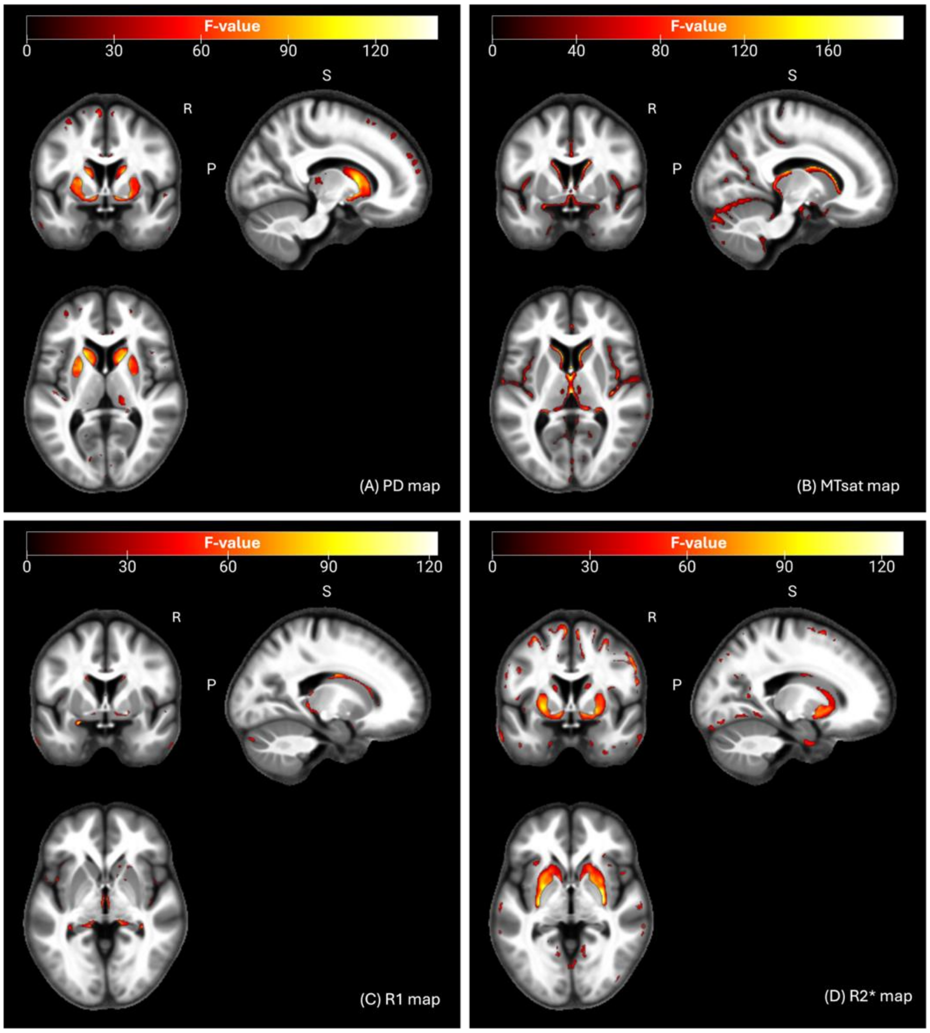


***Figure 1. Statistical parametric maps for the uGLMs at a corrected threshold of p<.05 FWER in the GM;*** *showing all the voxels with significant correlation with age, as detected by uGLMs for PD, MTsat, R1, and R2* maps. The F-tests were thresholded at p<0.05 FWER corrected at voxel-level. The SPMs were overlayed on the mean MTsat map for the cohort in the MNI space. Abbreviation: GLM, general linear model; uGLM, univariate GLM; GM, gray matter; FWER, family-wise error rate; SPM, statistical parametric map.*

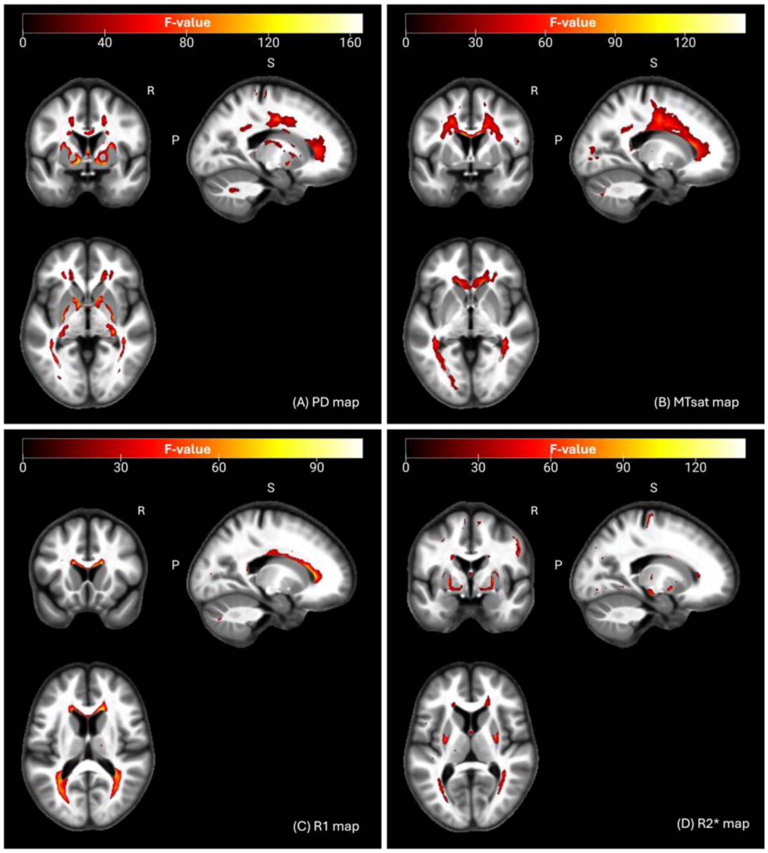


***Figure 2. Statistical parametric maps for the uGLMs at a corrected threshold of p<.05 FWER in the WM;*** *showing all the voxels with significant correlation with age, as detected by uGLMs for PD, MTsat, R1, and R2* maps. The F-tests were thresholded at p<0.05 FWER corrected at voxel-level. The SPMs were overlayed on the mean MTsat map for the cohort in the MNI space. Abbreviation: GLM, general linear model, uGLM, univariate GLM, mGLM, multivariate GLM, WM, white matter, FWER, family-wise error, SPM, statistical parametric map.*

#### 3.2 Canonical Vectors

*Table 1.* ***Canonical vector sizes for different modalities from the mGLM model****, representing the contribution of each modality in each voxel. The coordinates correspond to the peak voxels at the selected ROIs.*

| ROI name | Peak Coordinate (mm) | PD | MTsat | R1 | R2* |
|---|---|---|---|---|---|
| | | **Left** | | | |
| Putamen | (-28,-11,1) | 1.000 | 0.007 | 0.001 | 0.002 |
| Thalamus | (-6,-8,15) | 0.953 | 0.012 | 0.302 | 0.022 |
| Hippocampus | (-32,-39,-5) | 0.884 | 0.422 | 0.107 | 0.173 |
| Mid-Frontal g | (-27,-3,49) | 0.996 | 0.051 | 0.016 | 0.066 |
| Precentral | (-30,-24,64) | 0.832 | 0.137 | 0.247 | 0.478 |
| Heschl g | (-38,-22,12) | 0.946 | 0.024 | 0.322 | 0.027 |
| Supp-motor | (-7,-2,59) | 0.993 | 0.040 | 0.048 | 0.100 |
| Caudate | (-8,15,8) | 0.977 | 0.126 | 0.079 | 0.155 |
| Pallidum | (-28,-13,-4) | 1.000 | 0.006 | 0.001 | 0.001 |
| | | **Right** | | | |
| Putamen | (12,10,-3) | 0.922 | 0.285 | 0.079 | 0.248 |
| Thalamus | (10,-17,17) | 0.984 | 0.119 | 0.136 | 0.009 |
| Hippocampus | (33,-34,-3) | 0.951 | 0.027 | 0.302 | 0.063 |
| Mid-Frontal g | (32,1,52) | 0.980 | 0.158 | 0.010 | 0.125 |
| Precentral | (54,-1,42) | 0.894 | 0.397 | 0.084 | 0.190 |
| Heschl g | (37,-20,7) | 0.945 | 0.110 | 0.306 | 0.029 |
| Supp-motor | (5,-10,57) | 0.984 | 0.086 | 0.049 | 0.149 |
| Caudate | (10,16,10) | 0.985 | 0.075 | 0.141 | 0.063 |
| Pallidum | (11,8,-4) | 0.973 | 0.146 | 0.012 | 0.181 |

Key: MTsat, magnetization transfer saturation; PD, proton density; R2*, effective transverse relaxation rate; R1, longitudinal relaxation rate.

#### 3.3 Univariate and multivariate model evaluation

*Table 2.* ***Summary statistics for significant voxels in uGLMs and mGL in CV1.*** *"United" rows show the union of significant voxels in SPMs for all modalities. Univariate GLMs were thresholded at p<0.05 FWER corrected per tissue class (GM or WM). Abbreviation: GLM, general linear model; uGLM, univariate GLM; mGLM, multivariate GLM; GM, gray matter; WM, white matter.*

| Map name | #clusters | Cluster size range | #voxels |
|---|---|---|---|
| **uGLMs p<0.05 FWE corrected within GM** | | | |
| **MTsat** | 196 | 1-3177 | 9183 |
| **PD** | 75 | 1-2359 | 6322 |
| **R1** | 27 | 1-102 | 297 |
| **R2*** | 84 | 1-2117 | 5789 |
| **United** | 304 | 1-4053 | **18794** |
| **mGLM p<0.05 FWE corrected within GM** | | | |
| **All maps in mGLM** | 277 | 1-1302 | **11081** |
| **uGLMs p<0.05 FWE corrected within WM** | | | |
| **MTsat** | 32 | 1-8087 | 14026 |
| **PD** | 57 | 1-1384 | 8298 |
| **R1** | 23 | 1-3126 | 6268 |
| **R2*** | 36 | 1-446 | 2349 |
| **United** | 92 | 1-9519 | **22817** |
| **mGLM p<0.05 FWE corrected within WM** | | | |
| **All maps in mGLM** | 80 | 1-4079 | **13758** |

*Table 3.* ***Summary statistics for significant voxels in uGLMs and mGLM in CV1.*** *"United" rows show the union of significant voxels in SPMs for all modalities. Univariate GLMs were thresholded at p<0.0125=.05/4 FWER corrected, to account for the multiplicity of maps tested (4) per tissue class (GM or WM). The mGLM was thresholded at p<0.05 FWER corrected. Abbreviation: GLM, general linear model, uGLM, univariate GLM, mGLM, multivariate GLM, SPM, statistical parametric maps, GM, gray matter, WM, white matter.*

| Map name | #clusters | Cluster size range | #voxels |
|---|---|---|---|
| **uGLMs p<0.0125 FWE corrected within GM** | | | |
| **MTsat** | 163 | 1-1767 | 5872 |
| **PD** | 45 | 1-1046 | 3950 |
| **R1** | 14 | 1-69 | 142 |
| **R2*** | 43 | 1-1584 | 3787 |
| **United** | 214 | 1-2657 | **12141** |
| **mGLM p<0.05 FWE corrected within GM** | | | |
| **All maps in mGLM** | 277 | 1-1302 | **11081** |
| **uGLMs p<0.0125 FWE corrected within WM** | | | |
| **MTsat** | 33 | 1-4347 | 8352 |
| **PD** | 42 | 1-1027 | 5493 |
| **R1** | 23 | 1-2001 | 3798 |
| **R2*** | 24 | 1-354 | 1432 |
| **United** | 69 | 1-5237 | **14400** |
| **mGLM p<0.05 FWE corrected within WM** | | | |
| **All maps in mGLM** | 80 | 1-4079 | **13758** |

*Table 4.* ***Summary statistics for significant voxels in uGLMs and mGL in CV2.*** *"United" rows show the union of significant voxels in SPMs for all modalities. Univariate GLMs were thresholded at p<0.05 FWER corrected per tissue class (GM or WM). Abbreviation: GLM, general linear model; uGLM, univariate GLM; mGLM, multivariate GLM; GM, gray matter; WM, white matter.*

| Map name | #clusters | Cluster size range | #voxels |
|---|---|---|---|
| **uGLMs p<0.05 FWE corrected within GM** | | | |
| **MTsat** | 196 | 1-3887 | 11155 |
| **PD** | 42 | 1-3236 | 5861 |
| **R1** | 68 | 1-170 | 931 |
| **R2*** | 126 | 1-1006 | 6689 |
| **United** | 324 | 1-7321 | **21760** |
| **mGLM p<0.05 FWE corrected within GM** | | | |
| **All maps in mGLM** | 326 | 1-2125 | **13280** |
| **uGLMs p<0.05 FWE corrected within WM** | | | |
| **MTsat** | 36 | 1-2821 | 6973 |
| **PD** | 55 | 1-1116 | 10089 |
| **R1** | 30 | 1-390 | 861 |
| **R2*** | 54 | 1-1860 | 5364 |
| **United** | 111 | 1-3183 | **18875** |
| **mGLM p<0.05 FWE corrected within WM** | | | |
| **All maps in mGLM** | 148 | 1-2488 | **18042** |

*Table 5.* ***Summary statistics for significant voxels in uGLMs and mGLM in CV2.*** *"United" rows show the union of significant voxels in SPMs for all modalities. Univariate GLMs were thresholded at p<0.0125=.05/4 FWER corrected, to account for the multiplicity of maps tested (4) per tissue class (GM or WM). The mGLM was thresholded at p<0.05 FWER corrected. Abbreviation: GLM, general linear model, uGLM, univariate GLM, mGLM, multivariate GLM, SPM, statistical parametric maps, GM, gray matter, WM, white matter.*

| Map name | #clusters | Cluster size range | #voxels |
|---|---|---|---|
| **uGLMs p<0.0125 FWE corrected within GM** | | | |
| **MTsat** | 163 | 1-2080 | 7339 |
| **PD** | 24 | 1-2286 | 3925 |
| **R1** | 37 | 1-123 | 440 |
| **R2*** | 87 | 1-1006 | 3903 |
| **United** | 247 | 1-3224 | **13971** |
| **mGLM p<0.05 FWE corrected within GM** | | | |
| **All maps in mGLM** | 326 | 1-2125 | **13280** |
| **uGLMs p<0.0125 FWE corrected within WM** | | | |
| **MTsat** | 29 | 1-1616 | 3780 |
| **PD** | 55 | 1-1116 | 6780 |
| **R1** | 9 | 1-233 | 369 |
| **R2*** | 42 | 1-1199 | 3689 |
| **United** | 91 | 1-2021 | **12290** |
| **mGLM p<0.05 FWE corrected within WM** | | | |
| **All maps in mGLM** | 148 | 1-2488 | **18042** |

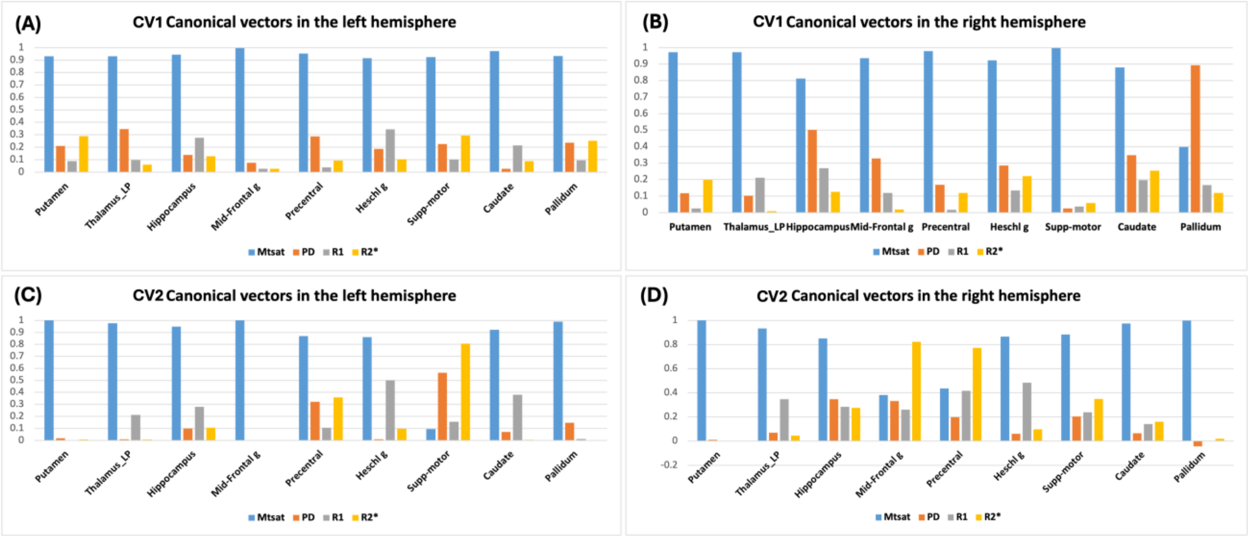


*Figure 3. Canonical Vectors for left and right hemispheres for CV1 and CV2*

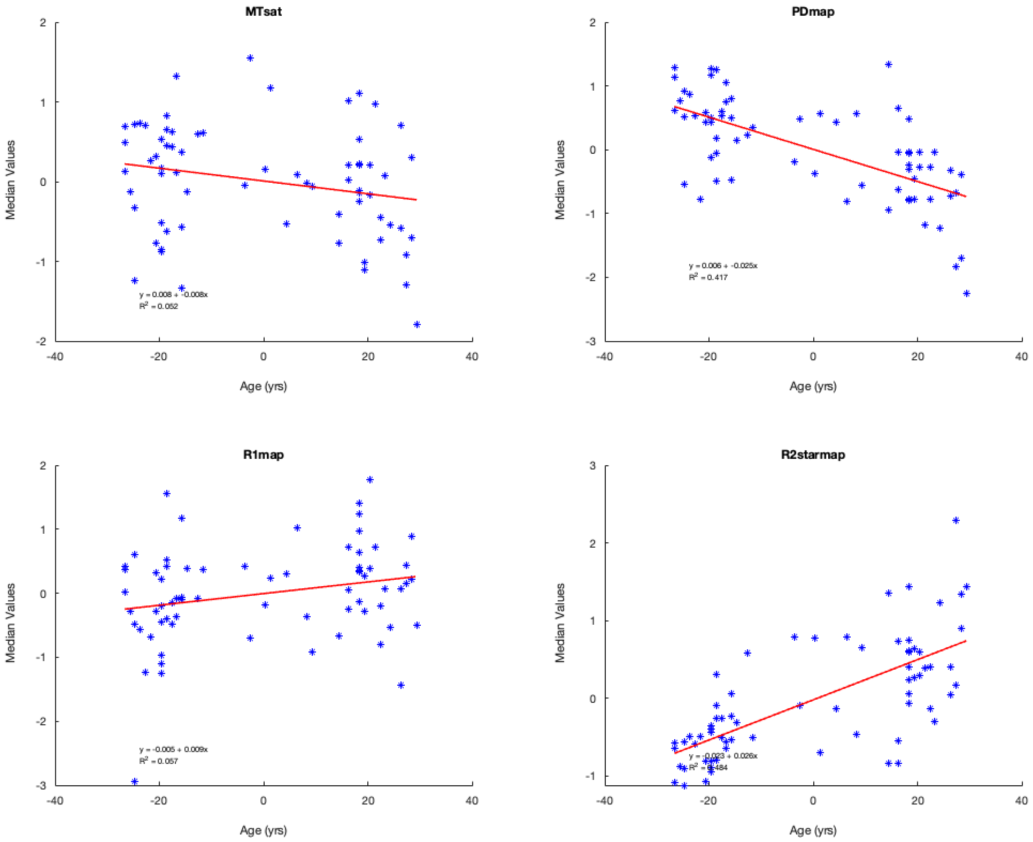


*Figure 4.* ***Median voxel values bilaterally in the Putamen in CV1.*** *The red lines depict the linear model fit. These data are shown for illustration purposes only and were not used for any additional analyses.*

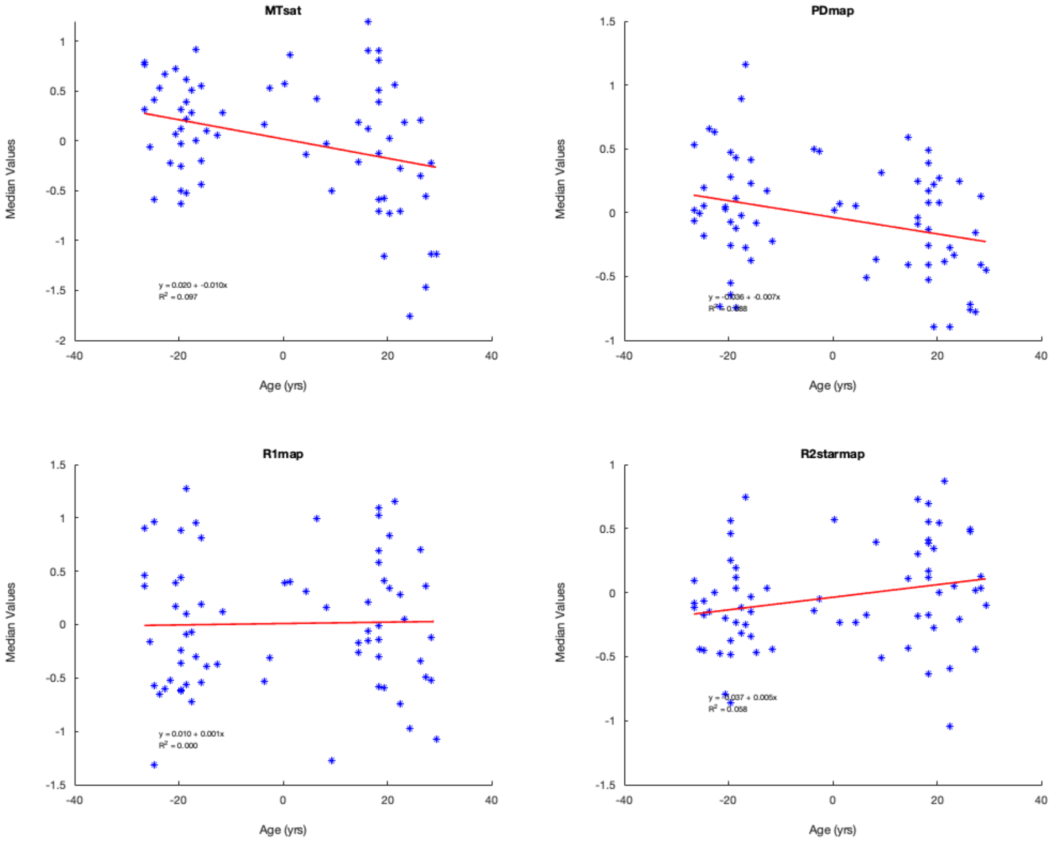


*Figure 5. **Median voxel values bilaterally in the Hippocampi in CV1.** The red lines depict the linear model fit. These data are shown for illustration purposes only and were not used for any additional analyses.*

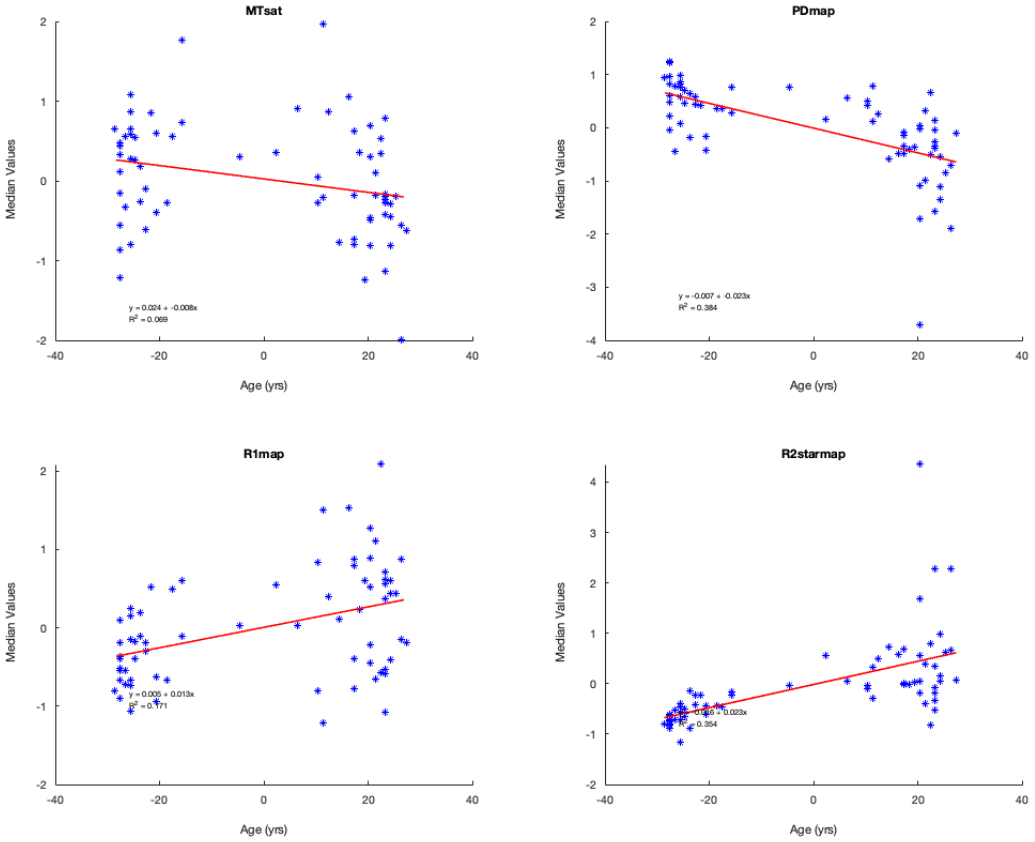


*Figure 6.* ***Median voxel values bilaterally in the Putamen in CV2.*** *The red lines depict the linear model fit. These data are shown for illustration purposes only and were not used for any additional analyses.*

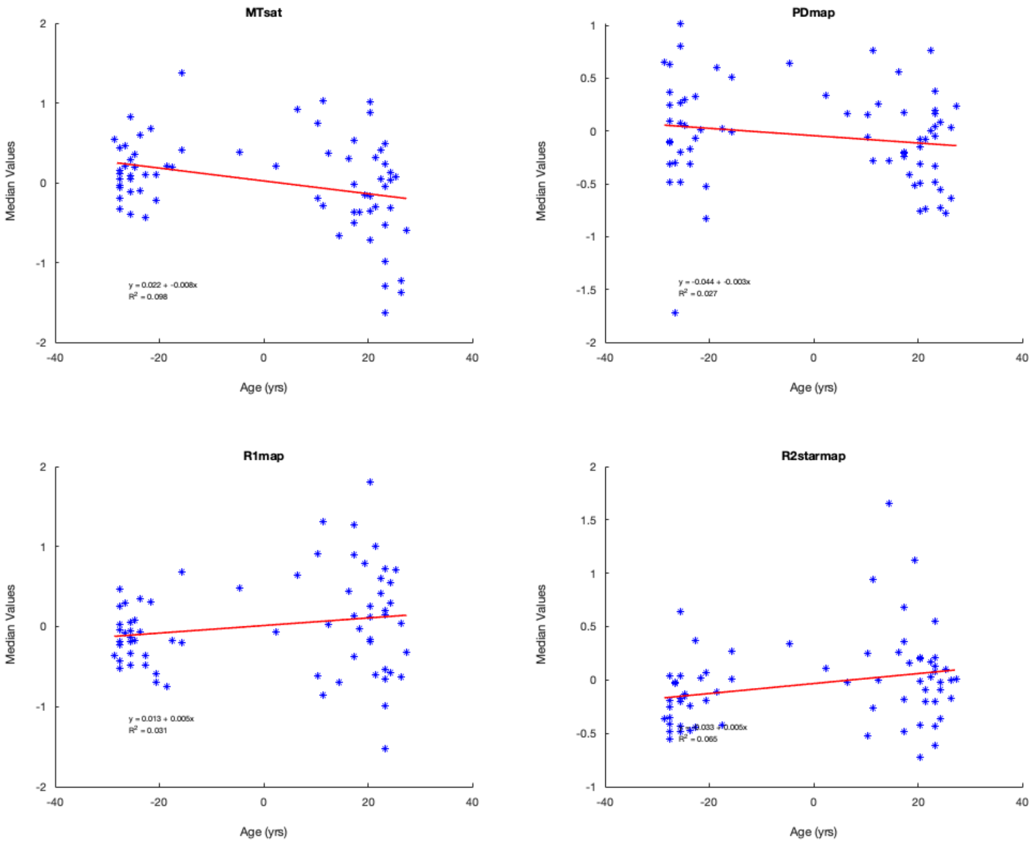


*Figure 7.* ***Median voxel values bilaterally in the Hippocmpi in CV2.*** *The red lines depict the linear model fit. These data are shown for illustration purposes only and were not used for any additional analyses.*

*Table 6.* ***Pearson Partial Correlations in CV1*** *on the median voxel values within different regions of interest.*

| | Pearson's r | p-value | Effect size (Fisher's z) | SE Effect size | | Pearson's r | p-value | Effect size (Fisher's z) | SE Effect size |
|---|---|---|---|---|---|---|---|---|---|
| **Caudate** | | | | | **Pallidum** | | | | |
| MTsat | -0.482* | < .001 | -0.526 | 0.123 | MTsat | -0.247* | .040 | -0.253 | 0.123 |
| PD map | -0.656* | < .001 | -0.786 | 0.123 | PD map | -0.645* | < .001 | -0.767 | 0.123 |
| R1 map | 0.059 | .628 | 0.059 | 0.123 | R1 map | 0.276* | .022 | 0.283 | 0.123 |
| R2s map | 0.508* | < .001 | 0.561 | 0.123 | R2s map | 0.684* | < .001 | 0.836 | 0.123 |
| **Cerebellum** | | | | | **Precentral Gyrus** | | | | |
| MTsat | -0.553* | <.001 | -0.622 | 0.123 | MTsat | -0.450* | < .001 | -0.485 | 0.123 |
| PD map | -0.403* | <.001 | -0.427 | 0.123 | PD map | -0.508* | < .001 | -0.560 | 0.123 |
| R1 map | -0.170 | 0.163 | -0.171 | 0.123 | R1 map | -0.028 | .822 | -0.028 | 0.123 |
| R2s map | 0.359* | 0.002 | 0.324 | 0.123 | R2s map | 0.319* | .007 | 0.331 | 0.123 |
| **Heschl Gyrus** | | | | | **Putamen** | | | | |
| MTsat | -0.543* | < .001 | -0.608 | 0.123 | MTsat | -0.229 | .058 | -0.233 | 0.123 |
| PD map | -0.222 | .066 | -0.226 | 0.123 | PD map | -0.646* | < .001 | -0.768 | 0.123 |
| R1 map | -0.330* | .006 | -0.343 | 0.123 | R1 map | 0.238* | .049 | 0.242 | 0.123 |
| R2s map | 0.067 | .586 | 0.067 | 0.123 | R2s map | 0.695* | < .001 | 0.858 | 0.123 |
| **Middle Frontal Gyrus** | | | | | **Superior motor cortex** | | | | |
| MTsat | -0.141 | .247 | -0.142 | 0.123 | MTsat | -0.205 | .091 | -0.208 | 0.123 |
| PD map | -0.576* | < .001 | -0.656 | 0.123 | PD map | -0.529* | < .001 | -0.589 | 0.123 |
| R1 map | 0.112 | .360 | 0.112 | 0.123 | R1 map | 0.119 | .329 | 0.120 | 0.123 |
| R2s map | 0.374** | .002 | 0.393 | 0.123 | R2s map | 0.555* | < .001 | 0.626 | 0.123 |
| **Hippocampus** | | | | | **Thalamus** | | | | |
| MTsat | -0.312* | .009 | -0.323 | 0.123 | MTsat | -0.517*** | < .001 | -0.572 | 0.123 |
| PD map | -0.296* | .013 | -0.305 | 0.123 | PD map | -0.405*** | < .001 | -0.430 | 0.123 |
| R1 map | 0.021 | .864 | 0.021 | 0.123 | R1 map | -0.240* | .047 | -0.245 | 0.123 |
| R2s map | 0.241* | .046 | 0.245 | 0.123 | R2s map | -0.240* | .047 | -0.245 | 0.123 |

*Note.* Conditioned on variables: Gender, TIV, TRIO. * p < .05

*Table 7.* ***Pearson Partial Correlations in CV2*** *on the median voxel values within different regions of interest.*

| | Pearson's r | p-value | Effect size (Fisher's z) | SE Effect size | | Pearson's r | p-value | Effect size (Fisher's z) | SE Effect size |
|---|---|---|---|---|---|---|---|---|---|
| **Caudate** | | | | | **Pallidum** | | | | |
| MTsat | -0.498* | < .001 | -0.546 | 0.123 | MTsat | -0.214 | .078 | -0.217 | 0.123 |
| PD map | -0.638* | < .001 | -0.755 | 0.123 | PD map | -0.727* | < .001 | -0.922 | 0.123 |
| R1 map | 0.071 | .560 | 0.071 | 0.123 | R1 map | 0.421* | < .001 | 0.449 | 0.123 |
| R2s map | 0.457* | < .001 | 0.494 | 0.123 | R2s map | 0.689* | < .001 | 0.846 | 0.123 |
| **Cerebellum** | | | | | **Precentral Gyrus** | | | | |
| MTsat | -0.586* | < .001 | -0.671 | 0.123 | MTsat | -0.345* | .004 | -0.359 | 0.123 |
| PD map | -0.250* | .039 | -0.255 | 0.123 | PD map | -0.414* | < .001 | -0.440 | 0.123 |
| R1 map | 0.074 | .544 | 0.074 | 0.123 | R1 map | 0.012 | .925 | 0.012 | 0.123 |
| R2s map | 0.444* | < .001 | 0.477 | 0.123 | R2s map | 0.541* | < .001 | 0.605 | 0.123 |
| **Heschl Gyrus** | | | | | **Putamen** | | | | |
| MTsat | -0.512* | < .001 | -0.565 | 0.123 | MTsat | -0.263* | .029 | -0.269 | 0.123 |
| PD map | -0.207 | .088 | -0.210 | 0.123 | PD map | -0.619* | < .001 | -0.724 | 0.123 |
| R1 map | -0.050 | .681 | -0.050 | 0.123 | R1 map | 0.413* | < .001 | 0.440 | 0.123 |
| R2s map | 0.358* | .003 | 0.374 | 0.123 | R2s map | 0.595* | < .001 | 0.685 | 0.123 |
| **Middle Frontal Gyrus** | | | | | **Superior motor cortex** | | | | |
| MTsat | -0.050 | .685 | -0.050 | 0.123 | MTsat | -0.042 | .730 | -0.042 | 0.123 |
| PD map | -0.448* | < .001 | -0.483 | 0.123 | PD map | -0.519* | < .001 | -0.575 | 0.123 |
| R1 map | 0.221 | .068 | 0.225 | 0.123 | R1 map | 0.214 | .077 | 0.218 | 0.123 |
| R2s map | 0.574* | < .001 | 0.653 | 0.123 | R2s map | 0.644* | < .001 | 0.764 | 0.123 |
| **Hippocampus** | | | | | **Thalamus** | | | | |
| MTsat | -0.314* | .009 | -0.325 | 0.123 | MTsat | -0.435* | < .001 | -0.467 | 0.123 |
| PD map | -0.164 | .179 | -0.165 | 0.123 | PD map | -0.233 | .054 | -0.237 | 0.123 |
| R1 map | 0.177 | .145 | 0.179 | 0.123 | R1 map | -0.172 | .159 | -0.173 | 0.123 |
| R2s map | 0.254* | .035 | 0.260 | 0.123 | R2s map | -0.122 | .320 | -0.122 | 0.123 |

*Note*. Conditioned on variables: Gender, TIV, TRIO. * p < .05
